# A myeloid program associated with COVID-19 severity is decreased by therapeutic blockade of IL-6 signaling

**DOI:** 10.1101/2022.11.07.22282049

**Authors:** Jason A. Hackney, Haridha Shivram, Jason Vander Heiden, Chris Overall, Luz Orozco, Xia Gao, Nathan West, Aditi Qamra, Diana Chang, Arindam Chakrabarti, David F. Choy, Alexis J. Combes, Tristan Courau, Gabriela K. Fragiadakis, Arjun Arkal Rao, Arja Ray, Jessica Tsui, Kenneth Hu, Nicholas F. Kuhn, Matthew F. Krummel, David J. Erle, Kirsten Kangelaris, Aartik Sarma, Zoe Lyon, Carolyn S. Calfee, Prescott G. Woodruff, Rajani Ghale, Eran Mick, Ashley Byrne, Beth Shoshana Zha, Charles Langelier, Carolyn M. Hendrickson, Monique G.P. van der Wijst, George C. Hartoularos, Tianna Grant, Raymund Bueno, David S. Lee, John R. Greenland, Yang Sun, Richard Perez, Anton Ogorodnikov, Alyssa Ward, Chun Jimmie Ye, UCSF COMET Consortium, Thiru Ramalingam, Jacqueline M. McBride, Fang Cai, Anastasia Teterina, Min Bao, Larry Tsai, Ivan O. Rosas, Aviv Regev, Sharookh B. Kapadia, Rebecca N. Bauer, Carrie M. Rosenberger

**Author notes:** Corresponding author: Carrie M Rosenberger, 1-650-467-9065, Additional UCSF COMET Consortium authors are listed in the acknowledgements. J.A. Hackney and H. Shivram are co-first authors.

## Abstract

Altered myeloid inflammation and lymphopenia are hallmarks of severe infections, including SARS-CoV-2. Here, we identified a gene program, defined by correlation with EN-RAGE (*S100A12*) gene expression, which was up-regulated in patient airway and blood myeloid cells. The EN-RAGE program was expressed in 7 cohorts and observed in patients with both COVID-19 and acute respiratory distress syndrome (ARDS) from other causes. This program was associated with greater clinical severity and predicted future mechanical ventilation and death. EN-RAGE^+^ myeloid cells express features consistent with suppressor cell functionality, with low HLA-DR and high PD-L1 surface expression and higher expression of T cell-suppressive genes. Sustained EN-RAGE signature expression in airway and blood myeloid cells correlated with clinical severity and increasing expression of T cell dysfunction markers, such as PD-1. IL-6 upregulated many of the severity-associated genes in the EN-RAGE gene program *in vitro*, along with potential mediators of T cell suppression, such as IL-10. Blockade of IL-6 signaling by tocilizumab in a placebo-controlled clinical trial led to rapid normalization of ENRAGE and T cell gene expression. This identifies IL-6 as a key driver of myeloid dysregulation associated with worse clinical outcomes in COVID-19 patients and provides insights into shared pathophysiological mechanisms in non-COVID-19 ARDS.

## Introduction

Altered myeloid cell expression states, including the accumulation of cells with hallmarks of myeloid-derived suppressor cells (MDSC), are consistent features in the blood of COVID-19 patients, and serve as a hallmark of severity (1–18). Monocytes and granulocytes associated with increased COVID-19 severity exhibit low expression of HLA-DR, and high expression of hallmark MDSC genes such as *S100A12* (EN-RAGE), and can impair T cell activation via contact-dependent (i.e., PD-L1) and soluble mechanisms, including IL-10, TGF-β, arginase 1, IDO-dependent tryptophan metabolism, and reactive oxygen and nitrogen species (reviewed in (19–22)). Presence of MDSCs in severe COVID-19 patients, and patients with other severe infections, correlates with reduced T cell numbers, and can impair T cell proliferation and IFN-γ production *ex vivo* (23–28). Reduced T cell proliferation and tissue sequestration can in turn contribute to the lymphopenia observed in COVID-19 patients with severe disease, which increases with the severity of respiratory failure and is prognostic for higher mortality(29). Thus, understanding the role of myeloid suppressor cells and the pathways leading to their dysregulation is of critical importance in COVID-19 and other infections.

The pathways driving myeloid inflammation and the mechanistic connection of maladaptive cellular programs to severe disease and response to drug interventions are not yet understood. IL-6 is a key regulator of inflammation and has been proposed as a potential driver of dysfunctional myeloid immune response in cancer, COVID-19, and other diseases (10, 12, 20, 26, 30–33). IL-6 treatment *in vitro* leads to differentiation of hematopoietic stem cells into CD14^+^ monocytes expressing a similar expression program to that observed in COVID-19 patient monocytes, including high expression of EN-RAGE and low expression of HLA-DR(10). Circulating IL-6 levels positively correlate with this severity-associated myeloid state in COVID-19 and other severe infections(10). It has been suggested that treatment with IL-6-blocking antibodies normalizes alterations in the myeloid compartment that are associated with disease severity(12), a hypothesis requiring placebo-controlled trials to test. Samples from hospitalized COVID-19 patients treated with tocilizumab (a monoclonal antibody against IL6R/Actemra) in a placebo-controlled study (COVACTA, n=438 hospitalized COVID-19 patients with hypoxemia randomized 2:1 tocilizumab:placebo) provided a unique opportunity to evaluate the role of IL-6 in shaping myeloid inflammation in patients(34). A meta-analysis by the World Health Organization concluded that IL-6 antagonists reduce 28-day mortality in patients hospitalized for COVID-19(35).

Here, we used single cell RNA-seq (scRNA-seq) of peripheral blood mononuclear cells (PBMCs) and bronchoalveolar lavage (BAL) fluid cells collected from COVID-19 patients, to identify a myeloid cell program shared across tissue compartments, defined by high expression of the EN-RAGE gene, among other inflammatory markers. Expression of this program was associated with more severe disease in our discovery cohorts. We replicated this finding using data from the COMET observational study, which includes sampling from endotracheal aspirates (ETA), whole blood, and PBMCs (2, 36, 37) of COVID-19 patients. Combined single cell profiling of RNA and cell surface proteins (CITE-seq) characterized the immunosuppressive expression program in EN-RAGE signature expressing cells and connected it to cell surface phenotypes of impaired myeloid antigen presentation and T cell dysfunction. The EN-RAGE expression program was associated with several measures of COVID-19 clinical severity and outcomes and was also observed in patients with acute respiratory distress syndrome (ARDS) from other causes, identifying a targetable pathway relevant to ARDS. Finally, blocking IL-6 signaling using tocilizumab in an interventional setting reduced expression of the EN-RAGE signature and normalized T cell numbers in COVID-19 patients from the COVACTA trial, providing mechanistic insight into the therapeutic response to COVID-19 patients to IL-6 blockade.

## Results

### A pan-myeloid EN-RAGE signature in blood and airway samples is associated with severe COVID-19

We used publicly available scRNA-seq data from COVID-19 patients to define a gene program that classifies a shared myeloid state across airway and blood samples. We seeded the program by the expression of the gene encoding EN-RAGE (*S100A12*), which has been implicated in several myeloid populations in the peripheral blood associated with COVID-19 severity(2, 10, 38, 39). EN-RAGE expression is upregulated by IL-6(40), is elevated in the airways of ARDS patients as well as other lung diseases(41), and serum levels correlate with COVID-19 severity(1, 4). We identified a set of 84 genes that co-vary (Pearson’s *r* > 0.5) with EN-RAGE expression across myeloid cells from bronchoalveolar lavage (BAL) fluid and PBMCs from COVID-19 patients(7, 11) (**Figure. 1A**; **Table S1**). This gene program showed strong coordinated expression across airway and blood samples and predominant expression in many myeloid cell types (monocytes, neutrophils, and macrophages), although dendritic cells (DCs) and non-classical monocytes showed lower overall expression (**Figure. S1**). Increased expression of the EN-RAGE program in neutrophil, monocyte and macrophage subsets was associated with greater clinical severity in both blood and airway samples (**Figure. 1B** and **Figure. S1C**).

**Figure 1.**
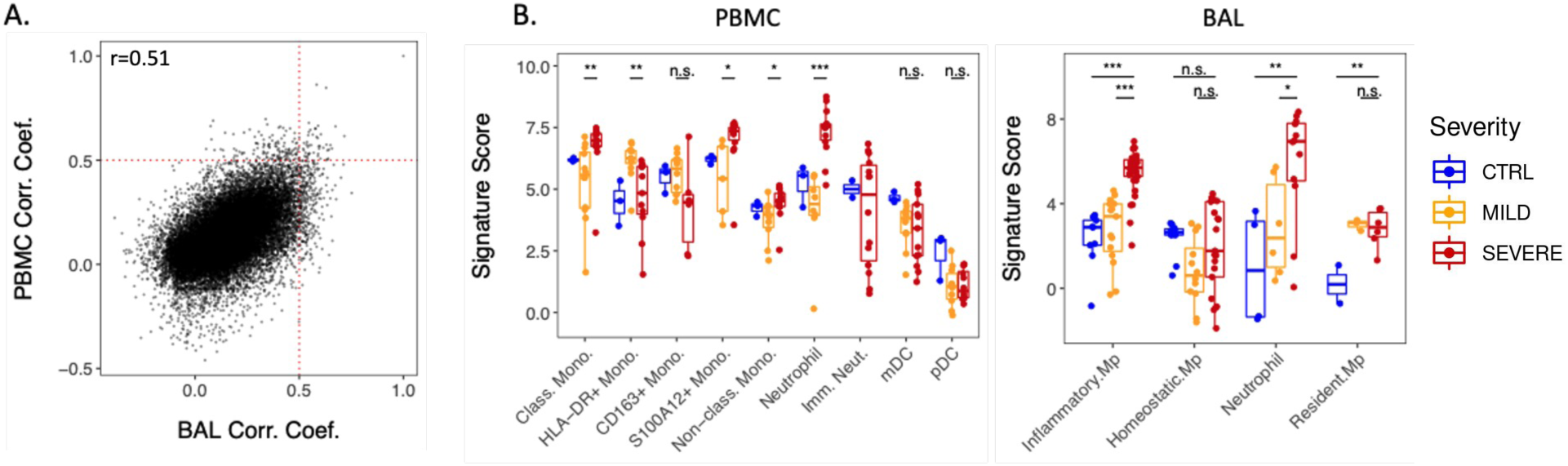
Identification of the severity-associated EN-RAGE myeloid signature in COVID-19 airway (BAL) and peripheral (PBMC) samples. A. Pairwise Pearson correlation between all genes and S100A12 in either PBMC(11) or BAL(7). B. Pseudo-bulk expression profiles of PBMC and BAL. Each point represents a patient. Blue=healthy (BAL n = 3; PBMC, n = 3), yellow=moderate/severe (BAL, n = 3; PBMC, n = 8; hospitalized -/+ supplemental O_2_), red=critical (BAL, n = 6; PBMC, n = 10; requiring mechanical ventilation), with severity defined within each dataset by the authors. Increased expression in severe patients in both tissues. Significance was tested using a t-test across the indicated groups (n.s. = p > 0.05, * = p < 0.05; ** = p < 0.01; *** = p < 0.001).

The EN-RAGE program scores correlated with the scores of a previously-defined MS1 gene set (9), which was associated with increased severity in COVID-19 and sepsis patients(9, 10) (Spearman’s ρ=0.64 and 0.95 in pseudobulk expression profiles of blood and airway monocytes and neutrophils (5 cohorts) and ρ=0.65 in whole lung myeloid cells (one cohort) (all p<0.0001)). Only seven genes (CLU, CYP1B1, LILRA5, NAMPT, S100A12, S100A8, VCAN) are shared between the 84 genes in the EN-RAGE program and the 23 genes in the MS1 signature identified in COVID-19 patients(10), which is a significant overlap (p-value=7e-14, hypergeometric test).

EN-RAGE program expression was more highly intercorrelated than the MS1 signature across sample types (**Figure. S2A)**. While MS1 had strong pairwise correlations in PBMC myeloid cells, many genes were no longer correlated when measured in whole blood (WB) or endotracheal aspirate (ETA) (**Figure. S2B-M)**. Because of the reduced performance of MS1 outside of PBMCs, we used the EN-RAGE program (denoted EN-RAGE^+^) in subsequent analyses to more specifically evaluate myeloid cells in bulk RNA-seq from whole blood and airway samples, which have greater cellular complexity than PBMC samples.

### EN-RAGE signature is associated with acute lung injury from diverse causes

We next hypothesized that the EN-RAGE myeloid program may also be a feature of non-COVID-19 lung injury. To test this hypothesis, we scored the EN-RAGE program in samples from the COMET observational cohort, where 75 patients with either COVID-19 or acute lung injury from other causes were followed longitudinally (**Table 1**, patient characteristics)(2, 36, 37). Whole blood (WB), PBMC, and endotracheal aspirates (ETA) were sampled and profiled by scRNA-Seq. This cohort offers rich clinical and molecular phenotyping to allow single-cell dissection of the connection between the airways and the blood at the mRNA and protein level, and how this relates to clinical outcomes. EN-RAGE signature expression was highest in monocytes, macrophages, and neutrophils, consistent with our previous analyses (**Figure 2** and **Figure S2**). EN-RAGE^+^ myeloid cells were present in the blood and airways in both COVID-19 and non-COVID-19 acute lung injury patients, highlighting the generality of this program (**Figure 2C**).

**Table 1.** Patient characteristics. NA = not available. COVACTA data are for subjects with blood RNA-seq data included in this manuscript. COVACTA ICU admission frequency is at time of baseline sampling.

|  | COMET |  |  | COVACTA |
| --- | --- | --- | --- | --- |
|  | All | SARS-CoV-2+ | SARS-CoV-2- |  |
| **n (%)** | 75 | 57 (76%) | 18 (24%) | 404 |
| **Age** median (IQR) | 54 (42, 69) | 48 (42, 66) | 66 (51, 76) | 63 (53-70) |
| **Sex** |  |  |  |  |
| Male n (%) | 52 (69%) | 43 (75%) | 9 (50%) | 284 (70%) |
| Female n (%) | 23 (31) | 14 (25%) | 9 (50%) | 120 (30%) |
| **Race** n (%) |  |  |  |  |
| White | 20 (27%) | 11 (19%) | 9 (50%) | 233 (58%) |
| Black/African American | 4 (5%) | 3 (5%) | 1 (6%) | 58 (14%) |
| Asian | 13 (17%) | 9 (16%) | 4 (22%) | 37 (9%) |
| Other/Mixed/Unknown | 38 (51%) | 34 (60%) | 4 (22%) | 76 (19%) |
| **Baseline NIH Ordinal Scale** n (%) |  |  |  |  |
| 3 No supplemental O2 | 14 (19%) | 12 (21%) | 2 (11%) | 15 (4%) |
| 4 Supplemental O2 | 25 (33%) | 18 (32%) | 7 (39%) | 109 (27%) |
| 5 Non-invasive/high flow O2 | 14 (19%) | 9 (16%) | 5 (28%) | 114 (28%) |
| 6 Mechanical ventilation (MV) | 3 (4%) | 3 (5%) | 0 (0%) | 58 (14%) |
| 7 MV + additional organ support | 19 (25%) | 15 (26%) | 4 (22%) | 107 (27%) |
| **SOFA maximal** median (IQR) | 4 (1, 10) | 4 (1, 11) | 4 (1, 9) | NA |
| **ICU admission** n (%) | 26 (35%) | 30 (53%) | 11 (61%) | 234 (58%) |
| **Mortality** n (%) | 10 (13%) | 6 (11%) | 4 (22%) | 83 (20%) |
| **ARDS AECC definition** n (%) | 38 (51%) | 26 (46%) | 12 (67%) | NA |
| **ARDS Berlin definition** n (%) | 23 (31%) | 18 (32%) | 5 (28%) | NA |
| **Hospitalization, days** median (IQR) | 8 (8, 18) | 11 (3, 20) | 5 (4, 7) | 22 (9-28) |
| **ICU duration, days** median (IQR) | 2 (0, 9) | 14 (4, 26) | 4 (2, 4) | 14 (0-27) |
| **Ventilator-free days** median (IQR) | 28 (18, 28) | 28 (16, 28) | 28 (26, 28) | 19 (0-28) |

**Figure 2.**
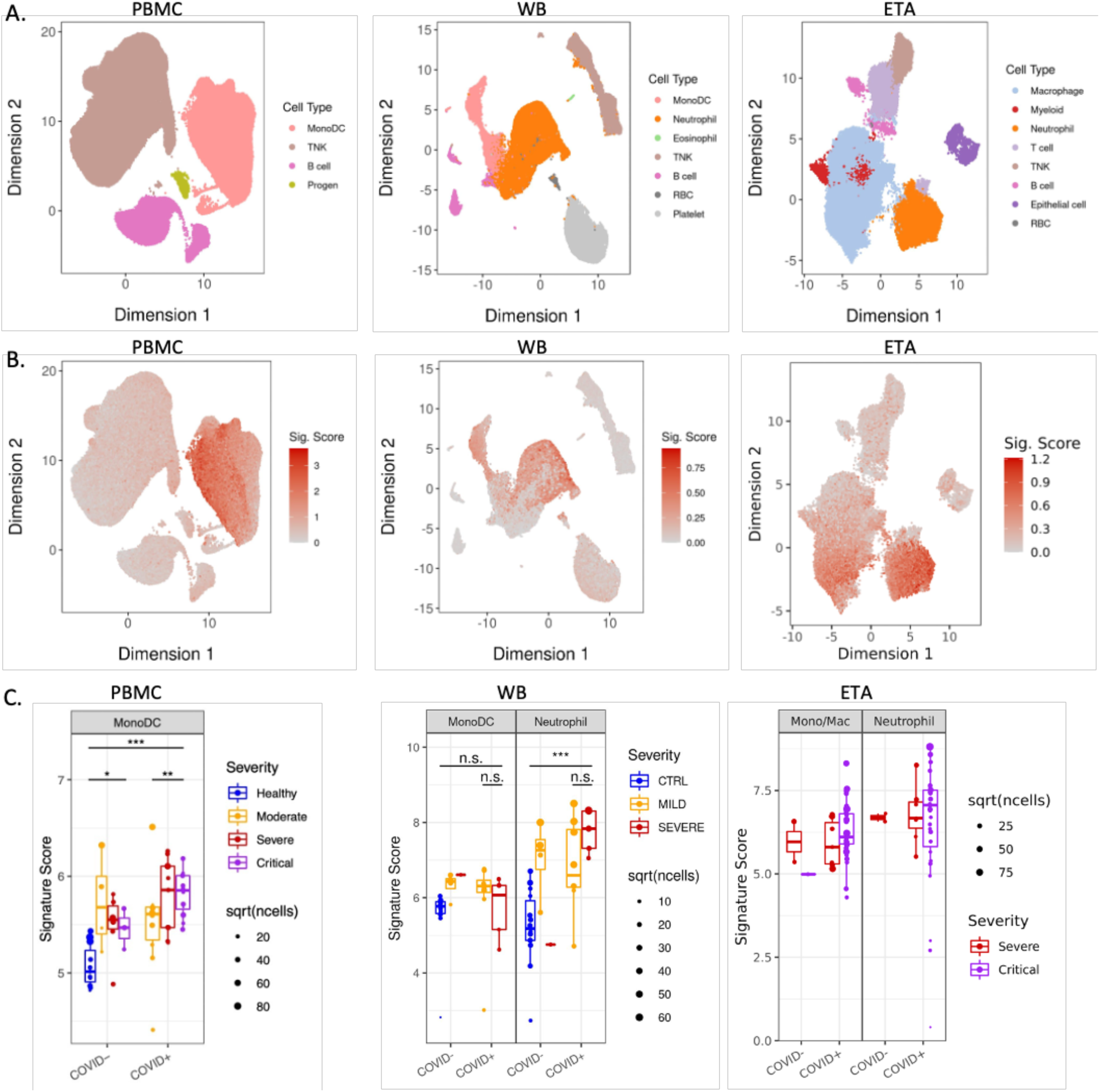
Replication of EN-RAGE severity association across sample types in both COVID-19 and non-COVID-19 acute lung injury (COMET cohort). A. UMAP plots with cell type annotations. Each point represents a single cell, colored by cell type. Each panel shows a different sample type, as indicated. PBMC: peripheral blood mononuclear cells, WB: whole blood, ETA:endotracheal aspirates. B. UMAP plots showing EN-RAGE signature score. Each point represented a single cell colored by the expression signature value. C. Pseudo-bulk expression profiles within myeloid cells. Each point represents the pseudo-bulk gene expression signature score for a cell type in a patient sample. PBMC severity: Moderate = no supplemental O_2_, severe = supplemental O_2_ and critical = mechanical ventilation. Whole blood severity: Mild/Moderate = 0 days on ventilator and no more than 1 day in ICU, Severe patients had ≥1 day on ventilator. ETA: Critical=VFDS=0 (ventilation for ≥28 days or death), severe ETA=VFDS>0. Significance was tested using a t-test across the indicated groups (n.s. = p > 0.05, ** = p < 0.01; *** = p < 0.001). Sample numbers per cohort: PBMC healthy, n = 11; PBMC COVID-moderate, n = 4; PBMC COVID-severe, n = 6; PBMC COVID-critical, n = 3; PBMC COVID+ moderate, n = 12; PBMC COVID+ severe, n = 10; PBMC COVID+ critical, n = 14. WB healthy, n = 14; WB COVID-mild, n = 4; WB COVID-severe, n = 1; WB COVID+ mild, n = 8; WB COVID+ severe, n = 5. ETA COVID-moderate/severe, n = 2; ETA COVID-critical, n = 1; ETA COVID+ moderate/severe, n = 5; ETA COVID+ critical, n = 8.

### EN-RAGE signature expression correlates with clinical severity and is prognostic for worse clinical outcomes

In COVID-19 patients, EN-RAGE^+^ myeloid cells were associated with increased clinical severity at presentation, as defined by the extent of respiratory support required at study enrollment, in monocytes (PBMC, **Figure 2C** and **Figure S1C**) and neutrophils (whole blood, **Figure 2C**). The severity association of monocyte ENRAGE score observed in PBMC was not observed in the smaller number of samples available from whole blood. An association with severity is not found in ETA samples, perhaps since this sample type is only obtained from critically ill patients on mechanical ventilation and so a milder severity group is lacking.

We next asked whether higher EN-RAGE signature expression predicts worse patient outcomes. In PBMC samples from COVID-19 and non-COVID-19 patients at COMET study enrollment, the EN-RAGE program score was associated not only with greater baseline clinical severity (NIH COVID-19 severity ordinal score) but also worse clinical outcomes (ICU admission p<0.01and maximal NIH ordinal scale: Spearman ρ=0.30 p=0.02) (**Figure 3A** and **Figure S3A-C**). However, these associations were no longer significant once baseline severity measures were considered in our analyses (p>0.5, data not shown), possibly because of the limited numbers of patients in this cohort and the increased risk of severe outcomes in patients presenting with greater severity. EN-RAGE program expression was not significantly associated with age (Spearman ρ=-0.02, p=0.88) nor with days from symptom onset to study enrollment (Spearman ρ=0.09, p=0.57). EN-RAGE score was higher in patients who presented with or later developed ARDS compared with those who did not by either AECC or Berlin diagnostic criteria (p<0.01 for AECC definition, p<0.05 for Berlin definition) (**Figure S3D-E**). ENRAGE score was higher in ARDS (AECC definition) resulting from SARS-CoV-2 infection or other etiology (p<0.05) (**Figure 3B**), but the difference in the smaller patient subgroups was not significant when using the more stringent Berlin ARDS definition (**Figure S3F**).

**Figure 3.**
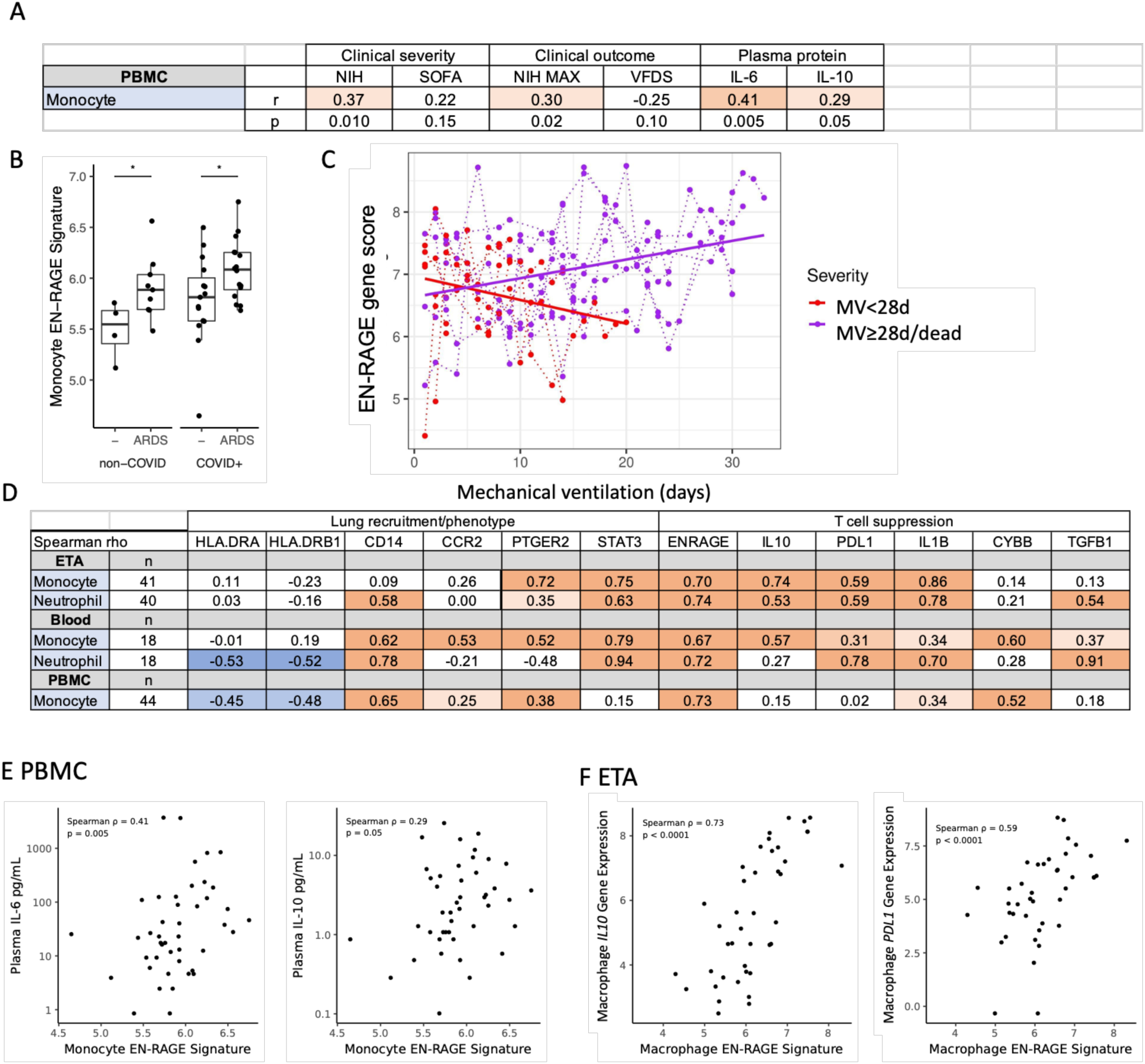
EN-RAGE signature expression correlates with disease severity and immunosuppressive gene expression in myeloid cells (COMET cohort). **A.** Spearman correlations between pseudo-bulk EN-RAGE signature score in PBMC monocytes and NIH ordinal severity score, maximal NIH severity score, SOFA organ failure score, and plasma IL-6 and IL-10 protein levels at study enrollment. **B.** PBMC monocyte EN-RAGE gene score is higher in patients who develop ARDS (AECC definition) in COVID-19 patients; n=46. Medians are indicated. * t-test p<0.05. **C.** Longitudinal changes in EN-RAGE signature in bulk ETA RNA-seq. Each point represents a patient sample from COVID-19 (n=16 patients, n=276 samples) and non-COVID (n=3 patients, n=6 samples) patients requiring mechanical ventilation. Samples from the same patient are linked by dotted lines. Points are colored by severity of disease. For illustrative purposes, linear regression trend lines for signature scores over time, grouped by severity level are shown as solid lines. Slopes were significantly different using a linear mixed model, p<0.05. MV=mechanical ventilation. **D.** Table of Spearman correlation coefficients between pseudo-bulk EN-RAGE signature score and genes encoding myeloid effector functions within monocyte or neutrophil populations across endotracheal aspirates (ETA), whole blood, and PBMCs from the COMET cohort. Positive correlations are shaded red and negative correlations shaded blue, with increasing darkness of shading indicating two tailed p values of p<0.05, p<0.01, and p<0.001. **E.** PBMC myeloid EN-RAGE gene score correlates with plasma IL-6 and IL-10 protein; n=46. **F.** Correlation of pseudo-bulk expression signature for EN-RAGE genes compared to pseudo-bulk expression values of IL-10 and PD-L1 in monocytes in COMET tracheal aspirate samples. Each point represents the expression value in a cell type in a single sample; n=40. Log_2_ gene expression, Spearman correlation coefficients and two tailed p values are shown.

Leveraging the availability of longitudinal samples from ventilated patients in the COMET cohort, there was a significant association between the temporal trajectory of airway EN-RAGE expression in each patient and patient outcomes (**Figure 3C**). To test this, we stratified patients into two groups by the number of ventilator-free days (VFD) and compared the slope of the regression lines of the two groups. Airway EN-RAGE expression decreased over time in survivors with fewest days of ventilation and increased in patients who died or had ≥28 days of ventilation (p<0.05 linear mixed model, ETA, **Figure 3C**). Worse clinical outcomes are accompanied by sustained elevated airway levels of EN-RAGE^+^ cells as well as higher baseline levels in the blood.

### EN-RAGE program expression in myeloid cells is associated with increased markers of immunosuppression in blood and airways

The EN-RAGE program score was also associated with expression of genes characteristic of MDSCs, suggesting one path through which EN-RAGE^+^ cells may contribute to clinical severity. Specifically, EN-RAGE program expression was correlated with metrics of suppressed myeloid and lymphoid states across another five COVID-19 cohorts(2, 5, 7, 11, 12, 36, 37), spanning ETA, BAL, lung, PBMC, and blood samples (complete results in **Table S2**). For example, in COMET PBMC monocytes, EN-RAGE program expression correlated with high *CD14* (Spearman ρ=0.65, p<0.001), *CCR2* (ρ=0.25, p<0.05) and *PTGER2* (ρ=0.38, p<0.001) (**Figure 3D**). *CCR2* and *PTGER2* are two receptors important for myeloid cell recruitment to the infected lung via *CCL2* and prostaglandins, respectively. EN-RAGE program expression correlated with low expression of MHC class II genes, suggesting reduced capacity for antigen presentation (*HLADRA:* ρ=-0.45 and *HLADRB1:* ρ=-0.48, p<0.001, **Figure 3D** and **Table S2**). In blood monocytes, there was a positive correlation with *STAT3*, a key transcription factor regulating MDSC gene expression (ρ=0.79, p<0.001, **Figure 3D**). MDSCs can suppress T cells using context-specific mechanisms across sites of infection or malignancy, and the mechanisms used can also differ depending on whether they originate from the monocytic or granulocytic lineage(13, 20). In blood monocytes, EN-RAGE program expression was positively correlated with expression of genes encoding effectors that can suppress T cells through reactive oxygen species (*CYBB/PHOX*) and prostaglandins (*PTGER2*), and *TGFβ1*, but inconsistent associations with arginase (*ARG1*) and tryptophan depletion (*IDO1*) across cohorts (**Figure 3D** and **Table S2**). Granulocytic EN-RAGE^+^ cells had similar associations as their monocytic counterparts, with notable differences including stronger correlations with *PDL1* and *TGFβ1* and little to no association with reactive oxygen species (*CYBB/PHOX*) and *PTGER2* compared with EN-RAGE^+^ monocytes (**Figure 3D** and **Table S2**). Consistent with MDSCs characterized in other infections and cancers(20, 22, 42), EN-RAGE^+^ cells expressed higher levels of multiple potential mediators of immunosuppression (*PDL1*, *CYBB/PHOX*, and *TGFβ1*) with some genes preferentially expressed by monocytic lineages (i.e. *CYBB/PHOX* ρ>0.4, **Table S2**) or granulocytic lineages (i.e. *TGFβ1* ρ>0.6, **Table S2**) (**Table S2**). EN-RAGE program expression in PBMC myeloid cells was modestly correlated with plasma protein levels of IL-6, a potential driver of MDSCs, and IL-10, a potential mediator of T cell suppression (Spearman ρ=0.41, p=0.005 and ρ=0.29, p=0.05, respectively; **Figure 3E**).

These associations were also largely observed in airways samples, suggesting an overall consistent phenotype in both the blood and the infected lung (**Figure 3D**). As in the blood, EN-RAGE program expression in monocytes and neutrophils was positively correlated with*, STAT3* in ETA samples but lacked the correlations with *CCR2* and *HLADR*, showed stronger correlations with *CCR5*, and inconsistent relationships with *CYBB* and *CD14* (**Figure 3D** and **Table S2**). EN-RAGE^+^ monocytes and neutrophils cells in lung and airway samples (ETA, BAL, and lung post-mortem autopsy tissue) had increased association with *PDL1*, *IL10, TGFβ, IDO,* and *IL1β*, which can suppress T cell function, compared to the blood (**Figure 3D,F** and **Table S2**). Peripheral blood myeloid cells expressed lower levels of *IL10* and *IL1β* and had weaker correlation with the EN-RAGE signature when transcripts were detected, illustrating the importance of sampling infected tissues and establishing blood correlates of tissue immune responses (**Figure 3D** and **Table S2**). Across four patient cohorts, airway EN-RAGE^+^ monocytes and neutrophils consistently expressed multiple markers of inflammation with autoregulatory functions that can be immunosuppressive, including *IL10, PDL1, TGFβ1, IL1β*, and *IDO* (**Table S2**).

### Characterization of EN-RAGE^+^ myeloid and T cell phenotypes

To connect the EN-RAGE expression program to cellular phenotypes, we used CITE-seq data from PBMCs in COMET to relate cell surface protein expression and EN-RAGE program expression. The EN-RAGE signature was most highly expressed in CD14^+^CD16^lo^HLA-DR^lo^ classical monocytes (cM) (**Figure 4A-B**), confirming the reduced antigen presentation capacity suggested by scRNA-seq data (**Table S2**). EN-RAGE signature scores on cMs positively correlated across patients with cM surface expression of 19 of the 188 CITE-Seq measured proteins (FDR<0.05, **Figure 4C**), including three markers characteristic of MDSCs: PD-L1 (CD274, Spearman ρ=0.43, FDR=0.008), podoplanin (PDPN, ρ=0.52, FDR=0.001), and CD38 (ρ=0.49, FDR=0.002), which is IL-6-inducible in tumors(43, 44). Conversely, EN-RAGE signature scores on cM were negatively correlated with 19 markers (FDR<0.05, **Figure 4C**), including HLA-DR (ρ=-0.73, FDR<0.0001), the T cell costimulatory protein CD40L (ρ=-0.36, FDR=0.03), the LFA subunit CD11A involved in trafficking and activation (ITGAL, ρ=-0.52, FDR=0.002), and two markers of granulocytic MDSCs (SIGLEC7: ρ=-0.52, FDR 0.001; CD244: ρ=-0.50, FDR=0.002) (**Figure 4C**).

**Figure 4.**
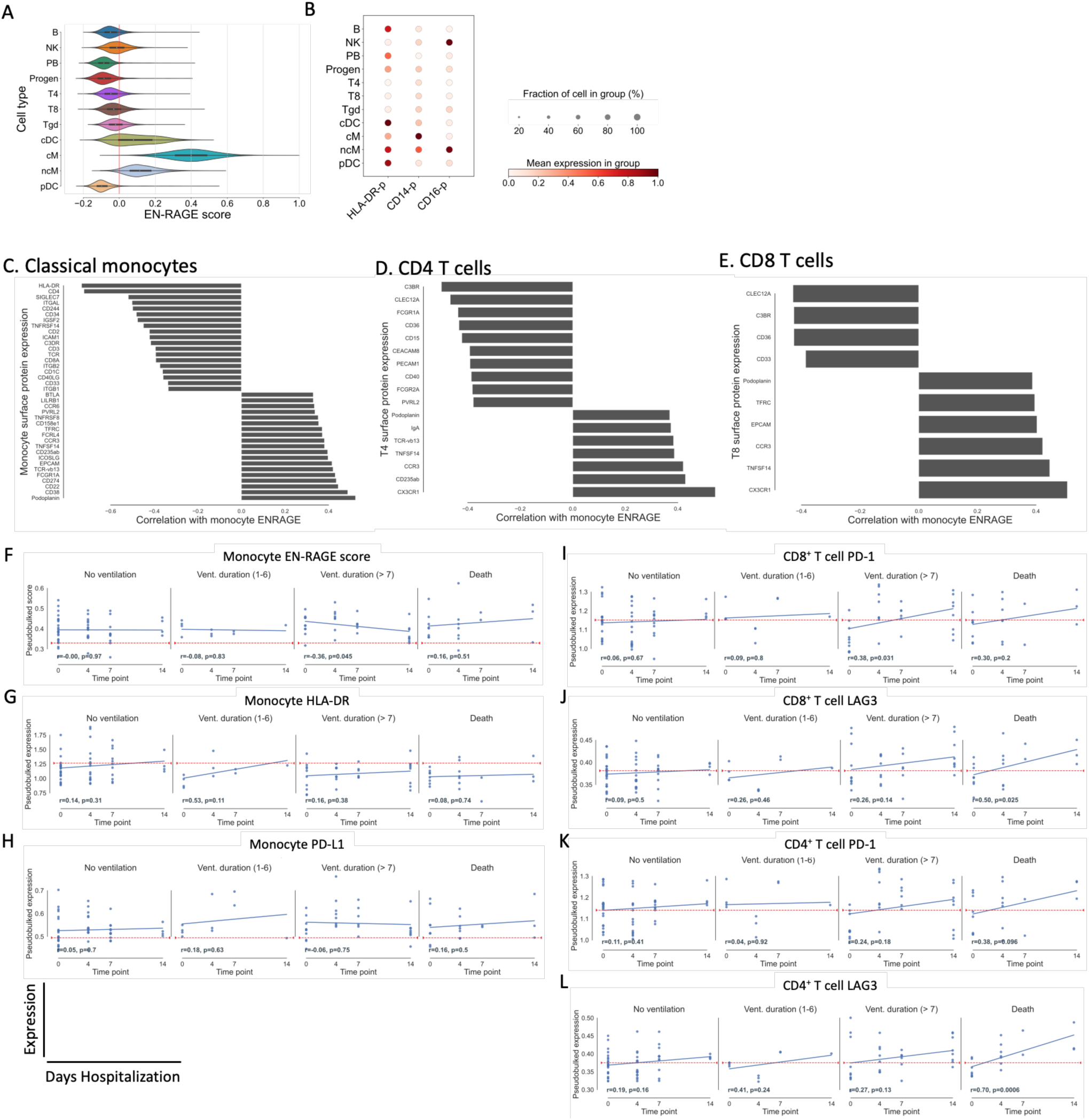
Characterization of myeloid and T cell immunosuppression phenotypes (COMET PBMC cohort). A. EN-RAGE gene set score is most highly expressed in classical monocytes (cM) in PBMC CITE-seq data. ncM=non-classical monocytes, progen=progenitor cells. Cells were defined by marker genes as previously described(37). B. CD14, CD16, and HLA-DR surface protein expression across cell lineages in PBMC CITE-seq data. C-E. Spearman correlations between EN-RAGE gene set expression in classical monocytes and protein expression on C. classical monocytes, D. CD4^+^ T cells and E. CD8^+^ cells; n=128 samples, including 11 healthy controls. FDR<0.05 for all correlations, panel of 188 proteins measured. F-L Pseudobulked surface protein expression in 128 PBMC samples from 60 patients over 14 days in patients grouped by clinical outcomes. Classical monocyte expression of F. EN-RAGE gene signature, G. HLA-DR protein, H. PD-L1 protein. CD8^+^ T cell expression of I. PD-1 and J. LAG3. CD4^+^ T cell expression of K. PD-1 and L. LAG3. Blue line denotes the linear regression trend for gene expression over time. Red line denotes expression level in healthy controls. Vent. duration = days of mechanical ventilation in survivors. n=128 samples from 60 patients (429, 505 cells). Pearson correlation coefficients (r) and p values are indicated.

EN-RAGE program scores in cMs also negatively correlated with the level of the activation marker CD40 on CD4^+^ T cells (**Figure 4D**), consistent with the negative correlation of the expression of ligand CD40L on cMs with their EN-RAGE program (**Figure 4C**). Conversely, EN-RAGE program scores in cMs were positively correlated with two proteins expressed by exhausted CD4^+^ and CD8^+^ T cells: podoplanin/PDPN (Spearman ρ=0.37, FDR=0.04) and TNFSF14/LIGHT (ρ=0.39, p=0.03) (**Figure 4D-E**). These data support the EN-RAGE program activity in cMs correlating with distinct CD8^+^ and CD4^+^ T lymphocytes activation states across patients.

Moreover, the dynamic changes in EN-RAGE program expression and markers of myeloid activation and T cell dysfunction were associated with clinical outcomes. Patients with PBMC CITE-seq data were categorized into 4 outcome groups based on survival and the duration of mechanical ventilation. In monocytes from patients with more severe outcomes compared with patients not requiring ventilation, HLA-DR trended lower than healthy (indicated by red line) and failed to recover to healthy levels over 14 days (**Figure 4F**), and trends of sustained higher PD-L1 and EN-RAGE score in some greater severity groups remained higher in ventilated patients (**Figure 4G-H**). On CD8^+^ and CD4^+^ T cells, markers of T cell dysfunction or exhaustion (PD-1, LAG3, TIGIT, CTLA4, and BTLA4) showed greater increases over 14 days in patients with worsening outcomes (>7 days mechanical ventilation and/or death) compared with non-ventilated patients (**Figure 4I-L** and **Figure S4**, Pearson correlation p<0.05, not significant for PD-1 on CD4^+^ T cells). Therefore, the EN-RAGE myeloid expression program correlates with markers of a suppressive cell surface phenotype on monocytes, and with reduced activation of CD4^+^ and CD8^+^ T cells, as indicated by increasing expression of T cell dysfunction markers over time (**Figure 4I-L** and **Figure S4**).

### IL-6 induces the EN-RAGE program in monocytes in vitro

Several lines of evidence led us to hypothesize that IL-6 can be a regulator of the EN-RAGE program. First, IL-6 treatment of HSPCs promotes upregulation of the MS1 gene signature in monocytes(10), which correlates with the EN-RAGE program (Spearman ρ≥20.64 in monocytes, p<0.0001 across 5 cohorts as described above). Moreover, blood monocyte EN-RAGE program expression correlates with plasma IL-6 protein levels, and with levels of monocyte *STAT3* mRNA, a transcription factor activated by IL-6 signaling (**Figure 3D-E** and **Table S2**). To test this hypothesis, we treated human primary monocytes with IL-6 *in vitro* followed by RNA-seq.

IL-6 treatment altered the expression of 36 of 84 EN-RAGE program genes (Benjamini-Hochberg FDR<0.05, **Figure 5A**), as well as of *IL10, IL1β, CYBB*, and *CCR2* (**Figure 5B**), all features of EN-RAGE^+^ monocytes in COVID-19 patients (**Figure 3D**). Using gene set enrichment analysis, we found that IL-6 could partially up-regulate the expression of EN-RAGE and MS1 signature genes (Benjamini-Hochberg adjusted p<0.001 and p<0.01, respectively; **Figure 5C**). Our data suggest that IL-6 is sufficient to upregulate the expression of many ENRAGE program genes in monocytes *in vitro*, including markers of a potentially T cell suppressive phenotype in patients.

**Figure 5.**
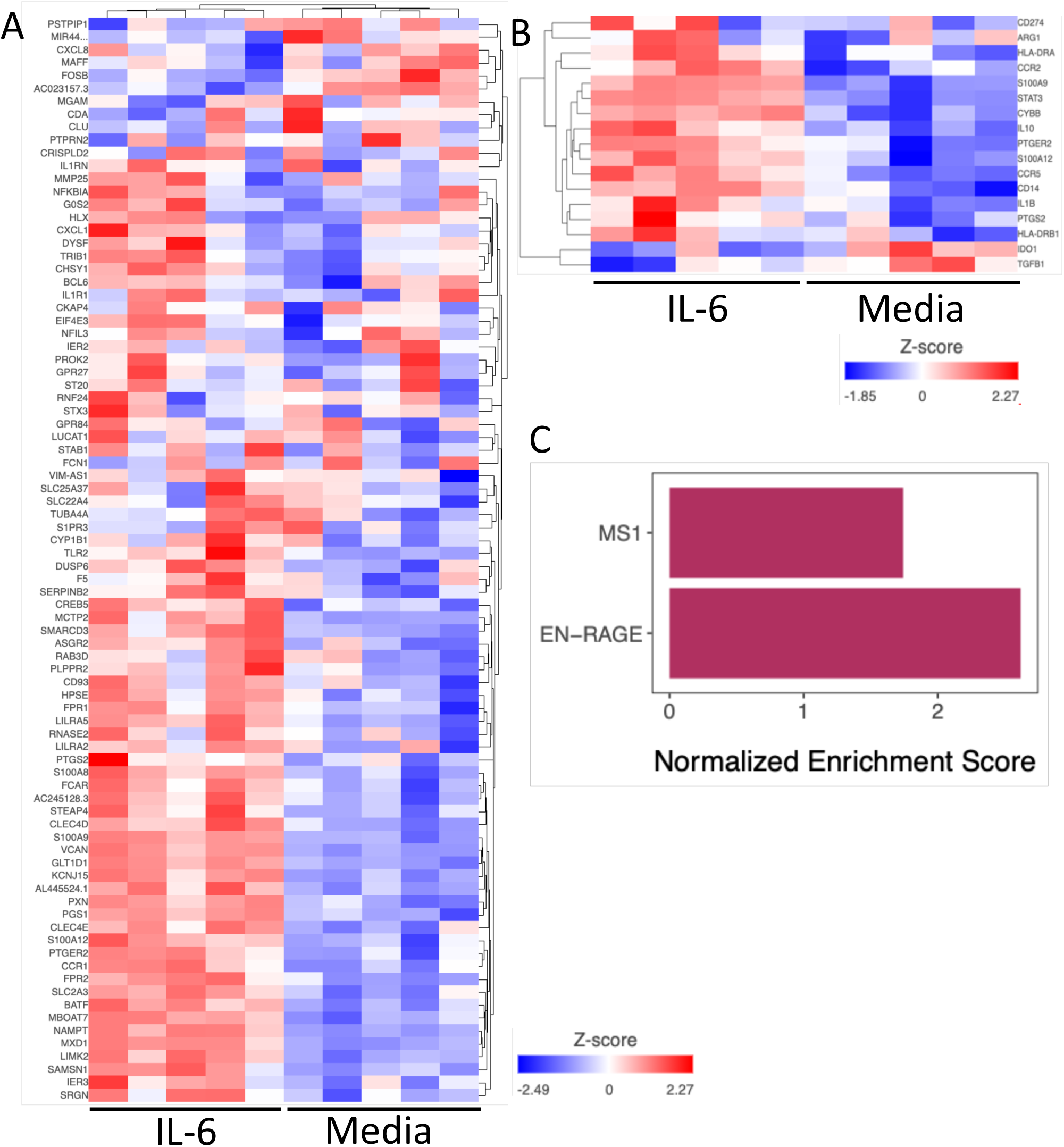
ENRAGE gene set enrichment analysis of monocytes treated with IL-6 *in vitro*. Monocytes stimulated with IL-6 for 24 hours compared with media, selected genes visualized as a heatmap using unsupervised clustering. A. ENRAGE signature genes. B. Genes associated with potential T cell suppressive functionality, from Supplemental Table 2. C. FGSEA analysis of EN-RAGE and MS1 signatures in IL-6 treated monocytes. Bars represent the normalized enrichment scores of how much each gene set is regulated by IL-6 treatment (BH adjusted p-value for MS1 < 0.01 and EN-RAGE < 0.001).

### The myeloid EN-RAGE program correlates with expression programs of suppressive myeloid cells and impaired T cells and with increased clinical severity in an interventional COVID-19 clinical trial

We next asked if blocking IL-6 signaling in patients correlated with a change in EN-RAGE program expression, leveraging data from COVACTA, a double-blind randomized clinical trial of tocilizumab (anti-IL6R/Actemra) in hospitalized COVID-19 patients with hypoxemia(34). Consistent with our findings in other cohorts, bulk RNA-seq expression profiles from whole blood collected at baseline from 438 patients showed higher normalized enrichment scores for the EN-RAGE program in patients requiring positive pressure ventilation at baseline compared with those who did not (**Figure 6A-B**, FGSEA Benjamini-Hochberg FDR < 0.05). The EN-RAGE gene set was also enriched in patients needing future mechanical ventilation or progressing to death even when controlling for the association with baseline severity (**Figure 6C-D**, FGSEA Benjamini-Hochberg FDR<0.05). The prognostic relationship of the ENRAGE signature with mortality was maintained even when adjusting for myeloid cell proportions in blood (**Figure S5A-E**; FDR<0.05, FGSEA). Moreover, lower expression of gene sets classifying CD8^+^ and CD4^+^ T lymphocytes (CIBERSORT(45)) was associated with worse clinical severity and outcomes (**Figure 6B-D**, (Benjamini-Hochberg FDR<0.05). Furthermore, consistent with COMET whole blood scRNA-seq, EN-RAGE program scores were positively correlated with expression of *PDL1, IL10,* and *IL1β* in whole blood at day 1 in COVACTA patients (**Figure 7A**), as well as with serum protein levels of EN-RAGE, IL-6, IL-10, IL-1β, and ARG1 (**Figure S7A**, t-test p<0.05). Examination of T cell genes that were associated with EN-RAGE program expression in scRNA-seq data (**Figure 7A**) revealed that high myeloid EN-RAGE program expression in COVACTA bulk RNA-seq was associated with low expression of T cell effectors (granzyme, perforin and lymphotoxin (*GZMB, GZMM, PRF1, LTA*)), cytotoxicity (*FASL*), and *IFNG*), activation markers (*KLRK1/NKG2D*) and dysfunction markers (*CTLA4, LAG3, CD160*). Many of these genes were expressed at a lower level in patients with more severe disease (*IFNG*, *FASLG, CTLA4, LAG3, TIGIT, TBX21/Tbet, XCL1*; **Figure 7B**, Benjamini-Hochberg FDR<0.05 and fold change < log_2_ -0.5). Higher EN-RAGE program expression was also correlated with lower lymphocytes (ρ=-0.5) and monocytes (ρ=-0.2) and increased neutrophils (ρ=0.5) (**Figure S7A**, all p<0.001). Thus, the COVACTA cohort shows similar features to those observed in COMET, including the expression of the EN-RAGE^+^ myeloid program and its correlation with greater clinical severity and worse clinical outcomes.

**Figure 6.**
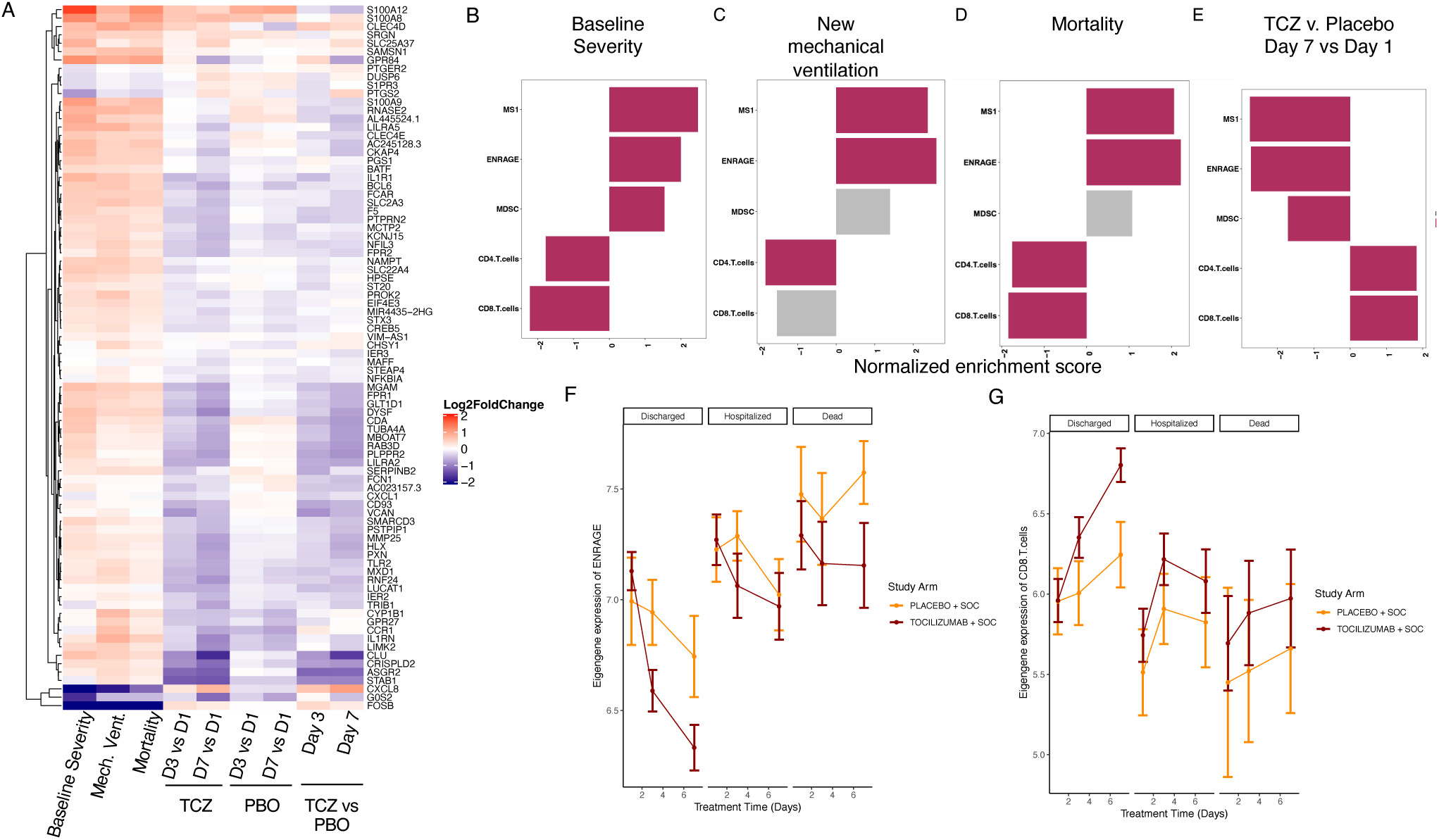
Severity-associated EN-RAGE gene set is associated with poor outcome and decreased by IL-6 blockade in COVID-19 patients (COVACTA cohort). **A.** Heatmap of EN-RAGE gene set associations with NIH ordinal scale severity at D1 (Severity NIH), need for new mechanical ventilation in patients not ventilated on D1 (Mech Vent), 28 day mortality (Death), treatment with tocilizumab or placebo at D3 or D7 relative to D1, and tocilizumab (TCZ) vs placebo at D7 relative to D1. **B-E.** Gene set enrichment analyses (GSEA) for gene sets associated with the myeloid cell states EN-RAGE, MS1(10), and MDSC(67) and T cells (CIBERSORT(45)). Normalized enrichment scores are shown, with red shading for t test p<0.05 and grey for p>0.05. TCZ = tocilizumab. For C and D, analyses were adjusted for baseline severity by incorporating baseline ordinal score as a covariate in our model. **F-G.** Tocilizumab treatment normalizes (F) EN-RAGE and (G) CD8^+^ T cell gene expression to healthy levels more rapidly than placebo in survivors. Only patients with measurements for all three time points are included. Lines represent the mean expression value for the gene set signature score across tocilizumab- or placebo-treated subjects. Error bars represent the 95% confidence interval around the mean. Patients are split into those that were discharged before 28, those that remained hospitalized, or subjects that died by day 28. Average signature scores are shown across the first 7 days of treatment. CTRL=healthy controls, SOC = standard of care drug therapy; significance testing was performed using t-test comparing each day to D1 by study arm * = p < 0.05, ** = p < 0.01, *** = p < 0.001, **** = p < 0.0001.

**Figure 7.**
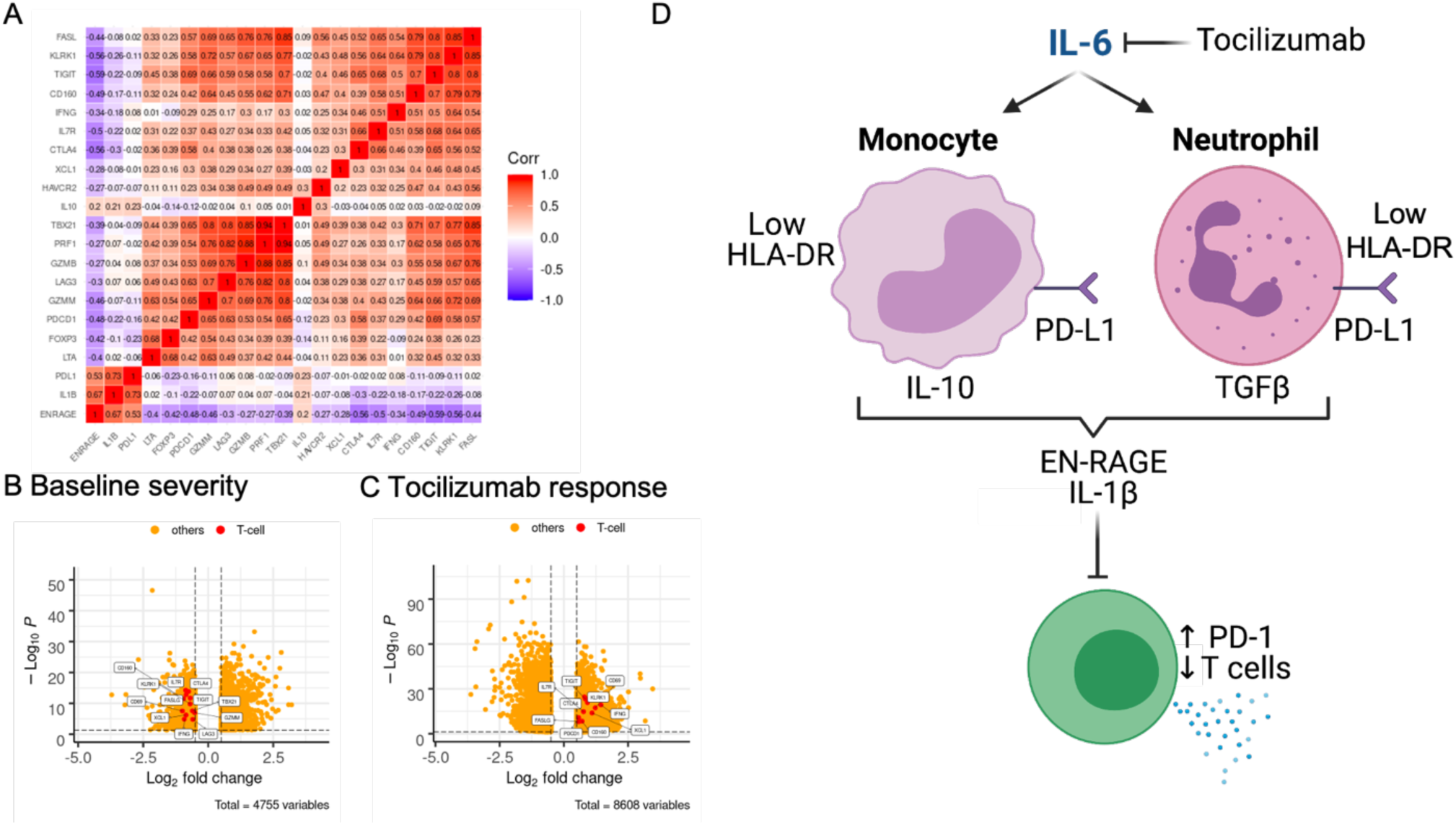
IL-6 blockade reduces potential T cell suppressive factors and normalizes T cells in COVID-19 patients (COVACTA cohort). A. EN-RAGE is positively correlated with IL-6-induced suppressive myeloid genes (*IL-10, IL-1b, PD-L1*) and inversely correlated with T cell genes (*FASL, LKRK1, TIGIT, CD160, IFNG, IL7R, CTLA4, XCL1, HAVCR2, TBX21, PRF1, GZMB, LAG3, GZMM, PDCD1, FOXP3*). Spearman correlation coefficients are shown for bulk whole blood gene expression. B-C. Volcano plots of reduced expression of T cell genes in A in patients with greater baseline severity (requiring positive pressure ventilation) versus not (B) and increased expression of T cell genes following 7 days treatment with tocilizumab (C). Dotted lines indicate absolute > 0.5 log_2_ fold change and FDR<0.05. D. Working model.

### IL-6 blockade decreases the myeloid EN-RAGE state and increases T cells in COVID-19 patients

Finally, to understand the role of IL-6 in patients, we examined the effect of tocilizumab treatment on the EN-RAGE myeloid state in COVID-19 patients and effect on T cells using longitudinal samples from the COVACTA study(14, 34). Blockade of IL-6 signaling reduced expression of many genes in the EN-RAGE program and increased T cell signature expression after 3 or 7 days of tocilizumab treatment compared with placebo (**Figure 6A, E**; Benjamini-Hochberg FDR<0.05). Consistent with the EN-RAGE program expression, MDSC and MS1 signatures were elevated in patients with worse baseline clinical severity and needing future mechanical ventilation or who died, and decreased following tocilizumab treatment (**Figure 6B-E**, Benjamini-Hochberg FDR<0.05). These enrichments were consistently maintained when adjusted for blood cell type composition (proportion of total leukocyte levels added as covariates to the DESeq2 model) (**Figure S5A-C**). The effect of tocilizumab on gene set enrichment was comparable between tertiles of baseline serum IL-6 protein levels (**Figure S5D-E**). EN-RAGE program gene expression was sustained over the first 7 days in patients with worse clinical outcomes at day 28 (hospitalized or non-survivor vs discharged, **Figure 6F**). The ENRAGE program was reduced following tocilizumab treatment more than in placebo treatment over the first 7 days, particularly in patients with better clinical outcomes (**Figure 6F** and **Figure S6A**). At the same time, CD8 T cell-associated gene expression rapidly increased after tocilizumab treatment, again particularly in patients that showed clinical improvement (**Figure 6F** and **Figure 7C**). Similar results were observed for the MS1 severity-associated myeloid and CIBERSORT CD4^+^ T cell gene sets (**Figure S6B, D**). This corresponded with a more rapid increase in blood lymphocytes and decrease in neutrophil cell counts in tocilizumab-treated patients who were discharged by 28 days compared with placebo (**Figure S6E-J**). When we compared the change between day 7 and 1 in EN-RAGE program expression with the change in measured blood cellularity, patients with a greater decrease in EN-RAGE had a greater increase in blood lymphocytes and monocytes and decreases in neutrophils (**Figure S7B-D**). This was more pronounced in patients treated with tocilizumab compared with placebo.

The reduction in EN-RAGE program following IL-6 blockade corresponded with normalization of potential mediators and correlates of T cell suppression. Patients with a greater decrease in EN-RAGE program expression from day 1 to 7 had greater decreases in serum EN-RAGE, IL-10, IL-1β, and ARG1 protein levels, as indicated by a positive slope of the correlation line (**Figure S7E-H**). IL-6R blockade with tocilizumab resulted in a greater decrease in ENRAGE program expression, as indicated by a greater offset in the regression in the tocilizumab-treated arm compared with placebo, and a greater decrease in this set of serum proteins, as indicated by more green vs red dots in the bottom left quadrant (**Figure S7E-H**). This correlated with increased expression of the T cell genes *IFNG, FASLG, CTLA4* and *XCL1* in tocilizumab-treated patients (**Figure 7C**). In sum, blockade of IL-6 signaling in COVID-19 patients rapidly normalizes the severity-associated myeloid and T cell states, identified using both patient scRNA-seq data and the IL-6 *in vitro* model, to levels in healthy individuals, which correlates with clinical improvement by 28 days.

## Discussion

While altered myeloid states are hallmarks of COVID-19 disease severity(1–18), the pathways driving maladaptive myeloid inflammation have not been clearly defined. This study supports a working model whereby IL-6 differentiates myeloid cells from both monocytic and granulocytic lineages to a suppressive phenotype characterized by low antigen presentation on HLA-DR and increased expression of multiple factors that can suppress T cells (IL-10, PD-L1, TGF-β1) (**Figure 7D**). We define a pan-myeloid ENRAGE program of coordinately-expressed genes in the airways and blood of COVID-19 patients that is prognostic for severe outcomes and is robust across 7 cohorts(2, 5–7, 11, 12, 14, 36, 37). EN-RAGE^+^ cells express multiple phenotypic hallmarks of MDSCs by cell surface protein analysis: decreased capacity for antigen presentation and co-stimulation through HLA-DR and CD40, and increased potential to suppress T cells through PD-L1. This was associated with sustained elevated expression of markers on T cells dysfunction such as PD-1 in patients with prolonged mechanical ventilation or who died. COVID-19 patients with higher EN-RAGE signature expression had a greater risk of future mechanical ventilation and mortality, and EN-RAGE^+^ myeloid cell impairment of optimal T cell-mediated immunity is one potential mechanism.

These data demonstrate the importance of IL-6 in altering immune cell phenotypes in COVID-19 patients and provide a potential mechanism for the therapeutic benefit of tocilizumab in patients hospitalized with COVID-19(35). Tocilizumab is approved for treating hospitalized adult COVID-19 patients requiring supplemental oxygen and receiving corticosteroids by the US Food and Drug Administration and the European Medicines Agency. IL-6 antagonists significantly reduced mortality compared with usual care in a large meta-analysis using data from 27 trials(35). While COVACTA did not meet the primary endpoint of improving clinical status on day 28, tocilizumab demonstrated clinically meaningful benefits, such as shortening hospital stay by 8 days, compared with the placebo arm(34). Therapeutic blockade of IL-6 signaling with tocilizumab attenuates expression of the EN-RAGE signature in blood cells and normalize T cell numbers, correlating with clinical improvement. IL-6 alone does not significantly induce the entire ENRAGE program, which is consistent with the partial IL-6R-dependent induction of the severity-associated MS1 program by sepsis patient serum using an *in vitro* system(10). The effect of IL-6R blockade in reducing the ENRAGE signature expression in patients was observed in survivors and not non-survivors (**Figure 6F**), suggesting other potential factors besides IL-6 that regulate expression of the full severity-associated transcriptional program.

COVID-19 patients with lymphopenia are at higher risk for worse clinical outcomes in the COVACTA cohort(29). In addition to being reduced in numbers, suppressed T cell phenotypes have been described in severe COVID-19 patients. Polyfunctional Th1 and Th17 CD4^+^ and CD8^+^ cell subsets are underrepresented in SARS-CoV-2 infection, with less proliferation and impaired IFN-γ and IL-2 secretion following restimulation *in vitro*(46, 47). T cells with increased expression of activation (OX40, CD69) and exhaustion (PD-1, TIGIT, TIM1) markers have been observed in some(38, 48) but not all cohorts(49) and functional exhaustion of CD8^+^ T cells has been reported(50–52). In single cell analysis of the COMET cohort, a phenotype of sustained elevated expression of PD-1, TIGIT, and LAG3 was observed on T cells in patients requiring longer mechanical ventilation and non-survivors. As tocilizumab increased the number of circulating T cells, the observed increase in markers of T cell function such as IFNγ and CD69 may result from increased T cell abundance rather than increased functionality per cell. Differential regulation of T cell genes suggests a potential effect of tocilizumab on T cell quality in addition to quantity. IL-6R blockade increases markers of early polyfunctional T cells (*IFNG, XCL1, CD69*) and recently activated/less exhausted cells (*IL7R*) while markers associated with broader spectrum of T cell functional states (i.e. cytotoxic T cell markers T-bet (*TBX21*), perforin (*PRF1*), and granzymes (*GZMM, GZMB*)) are less affected. However, there is limited ability to infer T cell functionality from bulk RNA-seq data. Future studies using multi-dimensional flow cytometry data will enable more precise definition of how IL-6 alters the T cell phenotype in COVID-19, both via myeloid activation and acting directly on T cells. NK cells are important antiviral effector cells that can be suppressed by MDSCs(19) and can express markers of dysfunction in COVID-19(49). A potential connection between EN-RAGE+ myeloid cells and NK cells or the suppressive regulatory T cell phenotype observed in severe COVID-19 patients(53) remains to be characterized.

Suppressive myeloid cells are one potential mechanism underlying impaired T cell immunity in severe COVID-19. MDSCs express mediators of T cell suppression in a context-dependent manner, depending on the type of infection or tumor(20, 22). MDSCs express multiple genes through which they may potentially suppress T cells. Moreover, differential correlations between the EN-RAGE signature and genes encoding immunosuppressive mediators were observed between cell lineage (granulocytic vs. monocytic) and compartment (blood vs. airways). MDSCs are often identified by low HLA-DR expression as a marker of impaired antigen presentation capacity(23, 26–28, 46). Interestingly, while some features of EN-RAGE^+^ cells were consistent between blood and airways (i.e. high PD-L1), the expression of other genes was tissue-dependent, and the low expression of HLA-DR in the blood was lost in airway samples. This is consistent with the observation that HLA-DR^lo^ MDSCs were not found in ETA airway samples by flow cytometry(26). These data suggest that EN-RAGE^+^ myeloid cells home to the infected lung and adopt a tissue-specific phenotype, with increasing expression of IL-10 and IDO and decreasing expression of CCR2 and CYBB, highlighting the importance of characterizing immune responses at the site of infection. The EN-RAGE^+^ myeloid state may underlie disease severity through additional mechanisms. For example, EN-RAGE^+^ cells express higher *IL-1β* and *TGF-β1* which can increase endothelial and epithelial cell permeability, respectively, and potential mechanisms whereby EN-RAGE^+^ cells could affect barrier integrity and edema in patients with acute lung injury remains to be explored(10, 54, 55).

This study connects immune cell phenotypes in the blood with the myeloid EN-RAGE state in the airways using paired quantification of mRNA and surface proteins at single cell resolution in PBMCs and ETA single cell transcriptomes from the COMET study and longitudinal blood mRNA expression data from the large Phase 3 COVACTA study. The inclusion of patients with acute lung injury from other causes in the COMET cohort reveals that EN-RAGE^+^ myeloid cells are relevant to insults in addition to SARS-CoV-2 infection, and identifies a targetable pathway relevant to ARDS. IL-6 predicts severe patient outcomes(29) and has been hypothesized to be a driver of myeloid differentiation in severe infections(10, 12). We show that tocilizumab treatment rapidly normalized myeloid and T cell states to healthy control levels, which was correlated with improved clinical outcomes. This study establishes the importance of IL-6 in driving differentiation of the severity-associated EN-RAGE^+^ myeloid state in patients. COVID-19 and cancer share risk factors, such as age and metabolic syndrome, and underlying immunobiology, such as MDSCs and T cell dysfunction, making it appealing to speculate that EN-RAGE^+^ myeloid cells may contribute to pathology in multiple diseases.

## Methods

### Reanalysis of published scRNA-seq data

Raw count data from Schulte-Schrepping, *et al.*(11) were downloaded from http://fastgenomics.org. For analysis of the PBMC data collected using 10x Genomics droplet-based capture, and whole blood collected using Rhapsody microwell-based capture, the raw count data was reprocessed using scripts written in the R programming language(56) and packages from the Bioconductor project (https://www.bioconductor.org). Briefly, counts were normalized using the computeLibraryFactors method from the scater R package(57) and dimensionality reduction was performed using PCA followed by UMAP projection. For calculating PCA, the top 5000 most variable genes were selected, accounting for overall expression level using the modelGeneVar method from the scran R package(58). The UMAP projection was calculated using the first 10 principal components. Graph-based clustering was performed using the Louvain algorithm on the same 10 principal components. Marker genes for each cluster were determined using pairwise t-tests, calculated using the pairwiseTTest method from the scran R package, and were used to manually assign broad cell type labels to each cluster. Clusters from the same broad cell type classification were merged. The myeloid populations (monocytes, DCs and neutrophils) were separately re-clustered using the same procedures: the 5000 most variable genes were selected, used for PCA calculation, graph-based clustering and UMAP projection. Marker genes were calculated for each cluster, and clusters were manually assigned using the marker gene lists.

Raw sequencing reads from Liao, *et al.*(7) were downloaded from SRA (PRJNA608742). Sequencing data were then processed using cellranger v4.0.0 (10x Genomics, Pleasanton, CA, USA) using GRCh38 as the reference genome, and gene models from GENCODE (v27) for assigning reads to genes. The emptyDrops method was used from the DropletUtils R package(59) to identify barcodes that corresponded to cellular droplets. Any barcode with fewer than 200 UMIs or 100 genes detected was removed. This allowed us to retain neutrophil-containing droplets, as these are mostly removed by default processing using cellranger. Droplets with high abundance of mitochondrial RNA (>10%) were removed. This yielded 90592 cells for further analysis. The same procedures described above were used to normalize, cluster and annotate cells into broad populations (macrophage, neutrophil, T cell, epithelial cell, B cell). The myeloid populations (macrophages and neutrophils) were selected and reclustered into more fine-grained populations using the same strategy as above.

Data from Silvin, et al.(12) were retrieved from the European Genome Archive (EGA) under accession number E-MTAB-9221 as FASTQ files. Sequencing data were processed as detailed above for PRJNA608742, using the same parameters for identifying cellular droplets and filtering droplets with high mitochondrial RNA abundance. Normalization, clustering and manual annotation was performed as described above. Count data from Delorey, et al.(5) were downloaded from the Broad Single Cell Portal using accession number SCP1052. We used the cell annotations as defined within that dataset, selecting cells labeled as Myeloid using the Cluster field defined by the authors. Data from Grant, et al.(6) were retrieved from the Gene Expression Omnibus (GEO) under accession number GSE155249. Normalization and clustering were performed as above. Annotations from the original publication were used to group cells into broad lineages: epithelial cells, B cells, T cells, dendritic cells, mast cells, macrophages and mixed myeloid cells.

### Pseudobulk expression profile calculation

The aggregateAcrossCells method from the scater R package(57) was used to calculate pseudobulk expression profiles for each cell population in a sample. This method uses the sum of raw counts to determine an estimate of the aggregate expression of that cell type in a sample. The TMM method from the edgeR R package(60) was used to normalize the pseudobulk count data within each dataset. Signature score calculation in pseudo-bulk data was performed as previously described(61) using the GSDecon package (https://github.com/JasonHackney/GSDecon).

### COMET PBMC CITE-seq analysis

The raw scRNA-seq counts were normalized using the ‘LogNormalized’ method implemented in the ‘NormalizeData’ function from the Seurat R package(62); with this method, the feature counts are divided by the total counts for each cell, multiplied by a scale factor of 10,000, and then natural-log transformed. The raw ADT counts were normalized using the ‘CLR’ method implemented in Seurat’s ‘NormalizeData’ function; with this method, a centered log ratio transformation is applied. The ENRAGE and MS1 gene set scores were calculated for each cell using the ‘score_genes’ function implemented in the scanpy Python package(63). All of the pseudobulk values for each cell population in a sample (i.e., sample + cell type)—that is, the mRNA expression, ADT expression, and gene set scores—were calculated as the average across all cells in that population. Correlations between gene set scores and ADT expression were on pseudobulk data and refer to a population of cells across patients.

### Identification of EN-RAGE gene expression induced by IL-6 in vitro

PBMCs were isolated from 50 mL of heparinized blood of four healthy donors by Ficoll-Paque. Primary human monocytes were purified from PBMC by Miltenyi Pan Monocyte Isolation Kit and cultured in RPMI with 10% heat-inactivated FBS, 10 mM HEPES and L-glutamine. The primary human monocytes were stimulated with 10 ng/mL IL-6 + 18.7 ng/mL IL6R for 24 hours, and RNA-seq performed on bulk cells. Unsupervised clustering using average linkage cluster difference and Euclidean point distance metrics were used to generate heat maps. RNA was isolated using the Qiagen RNeasy 96 kit (Qiagen catalog: 74182). Total RNA was quantified with Qubit RNA HS Assay Kit (Thermo Fisher Scientific catalog: Q32852) and quality was assessed using RNA ScreenTape on 4200 TapeStation (Agilent Technologies catalog: 5067-5576). cDNA library was generated from 2 nanograms of total RNA using Smart-Seq V4 Ultra Low Input RNA Kit (Takara catalog: 634894). 150 picograms of cDNA was used to make sequencing libraries by Nextera XT DNA Sample Preparation Kit (Illumina catalog: FC-131-1024). Libraries were quantified with Qubit dsDNA HS Assay Kit (Thermo Fisher Scientific catalog: Q32851) and the average library size was determined using D1000 ScreenTape on 4200 TapeStation (Agilent Technologies catalog: 5067-5582). Libraries were pooled and sequenced on the Illumina NovaSeq 6000 to generate 30 million single-end 50 base pair reads for each sample. Sequencing reads were filtered and aligned using HTSeqGenie v4.4.2(64). GSNAP v2013-11-01 was used for alignment, through the HTSeqGenie wrapper, against the GENCODE 27 Basic gene model on the human genome assembly GRCh38. Only reads with unique genomic alignments were analyzed. Normalized CPM (Counts Per Million) were used as a normalized measure of gene expression, calculated using method provided in edgeR(60).

### COMET cohort

The COVID-19 Multi-Phenotyping for Effective Therapies (COMET) cohort collected PBMCs, whole blood, ETA and plasma longitudinally from hospitalized patients presenting with COVID-19 symptoms. This study included 75 patients with samples collected in 2020, of whom 57 were positive (76%) for SARS-CoV-2, along with 11 healthy controls(2, 36, 37). Table 1 summarizes the patient characteristics. All-cause mortality occurred within 30 days for 9 of 10 subjects. Ventilator-free days (VFDS) were assessed at D28, with fatal cases assigned 0. NIH COVID-19 ordinal severity score and Sequential Organ Failure Assessment (SOFA) scores were assessed at study enrollment (Day 0), when baseline samples were collected, with a 9 day median time from symptom onset (4-13 IQR). Severity groups were defined as follows. PBMC: Moderate = no supplemental O_2_, severe = supplemental O_2_ and critical = mechanical ventilation. Whole blood: Mild/Moderate = 0 days on ventilator and no more than 1 day in ICU, Severe patients had ≥1 day on ventilator. ETA: Critical=VFDS=0 (ventilation for ≥28 days or death), severe ETA=VFDS>0.

PBMCs were isolated and single cell RNA sequencing and Cellular Indexing of Transcriptomes and Epitopes by Sequencing (CITE-seq) data was generated as previously described(37), with cell types defined by marker genes. The complete protocol is available on protocols.io (https://www.protocols.io/view/10x-citeseq-protocol-covid-19-patient-samples-tetr-bqnqmvdw). Data was generated for 188 unique cell surface antigens. CITE-seq data was included from 128 samples collected at day 0, day 7 and/or day 14 from 60 patients. Single cell RNA-seq data generated from PBMC collected at day 0 from 49 patients, whole blood collected at day 0 for 18 patients, and 41 ETA samples collected longitudinally from 16 patients were included in this study. Bulk gene expression data was generated from 182 samples collected longitudinally from 19 patients. Plasma cytokine and paired ETA scRNA-seq data was available for only 7 samples, precluding correlation analyses.

### COVACTA tocilizumab clinical trial

A total of 438 hospitalized COVID-19 patients were randomized 2:1 for anti-interleukin-6 receptor antibody, tocilizumab, or placebo and included in the modified intention to treat population (tocilizumab: 294, placebo: 144). Hospitalized patients were ≥18 years of age with COVID-19 pneumonia confirmed by a positive SARS-CoV-2 PCR test and evidenced by x-ray or computed tomography (CT) scan. Eligible patients had a blood oxygen saturation of ≤93% or partial pressure of oxygen/fraction of inspired oxygen of <300 mm/Hg. Details of the COVACTA study design have been published (Clinical trials.gov NCT04320615)(34). Population demographics for the 404 patients with available blood RNA-seq data are described in Table 1.

Serum IL-6 was quantified using a validated in vitro diagnostic method (Roche Cobas; Roche Diagnostics, Indianapolis, IN) at central laboratories (PPD). Complete blood counts were measured using standard clinical chemistry and haematology methods available at local hospital laboratories. IFNγ and IL10 were measured by qualified immunoassays (Simpleplex, ProteinSimple, San Jose, CA, USA) at central laboratories (Covance). The Olink Explore platform was used to measure 1472 serum proteins (Olink, Uppsala, Sweden). RNA was isolated from blood PaxGene (Qiagen, Hilden Germany) samples by Q2 Solutions. 1.25 ug of RNA was used for generating sequencing libraries with the TruSeq® Stranded mRNA Library Prep kit (Illumina, San Diego, CA, USA). The libraries were sequenced by Illumina NovaSeq by 50 bp single-end reads at a read depth of 50 million reads per sample.

For the time point comparisons (D1 to D7) patients were subset to those subjects with measurements at both time points before differential expression analysis with DESeq2. Unfiltered DESeq2 outputs were ranked by log2 fold change and then the FGSEA Bioconductor package was used to calculate enrichment scores. We used the ggplot2 package to generate visualizations. T cell genes with a relationship to EN-RAGE state in COMET scRNA-seq data were examined in COVACTA, and limited to the subset of T cell suppressive genes that were predominantly expressed by T cells in COMET whole blood transcriptome data to permit analysis of bulk gene expression data. Computational methods are described in more detail elsewhere(14).

### COVID-19 clinical severity

Within the COVACTA clinical trial, the NIH COVID-19 ordinal severity scale was used to assess disease severity: 3=hospitalized not requiring supplemental O_2_, 4= supplemental O_2_, 5= non-invasive/high flow O_2_, 6= mechanical ventilation, 7= mechanical ventilation + additional organ support (eg, vasopressors, renal replacement therapy, ECMO), 8=death. In the COMET cohort of PBMC samples, moderate = no supplemental O_2_ (NIH ordinal scale 3-4), severe = supplemental O_2_ (5–6) and critical = mechanical ventilation (7). The maximal NIH ordinal severity scale recorded during hospitalization was calculated for each patient in COMET. Ventilator-free days were calculated over 28 days, with fatal patients scored as 0. ARDS was diagnosed using the American-European Consensus Conference (AECC) definition(65) or Berlin definition(66). The sequential organ failure assessment (SOFA) score was calculated at study enrollment.

### Statistical analysis

Gene expression was log_2_ normalized. Patient demographics are given as median (IQR) and frequency (%). Medians and first and third quartile ranges are shown in box and whisker plots and medians are shown on dot plots. Longitudinal COVACTA line plots show means and 95% confidence intervals. Two-sided unpaired t-tests were used to compare gene signature scores between cell types in pseudobulk scRNA-seq data. Student’s t-test was used to calculate p values for comparisons of EN-RAGE gene set scores and clinical severity. A mixed linear model was used to compare slopes of longitudinal EN-RAGE gene set expression between severity groups. Spearman correlations and two-tailed p values were calculated to examine relationships between biomarkers and clinical severity and EN-RAGE gene set vs. myeloid gene expression. False discovery rates were calculated for differential expression analysis of bulk RNAseq data (accounting for transcriptome-wide analysis of x genes), GSEA (accounting for the x gene sets analyzed), and COMET PBMC CITE-seq data (accounting for 188 measured proteins), using the Benjamini-Hochberg method. Biorenderer was used to generate some Figures.

### Study approval

The COMET study was approved by the Institutional Review Board: UCSF Human Research Protection Program (HRPP) IRB# 20-30497 and informed consent was obtained for patients. COVACTA was conducted in accordance with Good Clinical Practice guidelines of the International Council for Harmonisation E6 and the Declaration of Helsinki or local laws and regulations, whichever afforded greater protection. Informed consent was obtained from the patient or their legally authorized representative prior to participation. The studies were approved by institutional review boards or ethics committees at each site.

## Data Availability

The RNAseq and proteomics data that support the findings of this study are available in the public online respositories Gene Expression Omnibus (GEO), SRA, and the European Genome-Phenome Archive (EGA). COMET: GEO GSE163668 (whole blood), GSE163426 (tracheal aspirate), GSE168453 (PBMC). Schulte-Schrepping, et al.: http://fastgenomics.org. Liao, et al.: SRA PRJNA608742. Silvin, et al.: EGA E-MTAB-9221. Delorey, et al.: Broad Single Cell Portal accession number SCP1052. Grant, et al.: GEO GSE155249. COVACTA RNA-seq, proteomics, and clinical metadata: EGA; accession number EGAS00001006688 (available to qualified researchers upon request; https://ega-archive.org/). Individual patient level clinical data for the COVACTA study is available through the clinical study data request platform (https://vivli.org/).

## Code availability

No new algorithms were developed for this manuscript. Most of the analyses performed in this study used published packages mentioned in the Methods, with the exception of the EnhancedVolcano (https://github.com/kevinblighe/EnhancedVolcano) package used to generate volcano plots. All code generated for analysis is available from the authors upon request.

## Author Contributions

C.M.R., J.A.H, and H.S. conceived of and designed the overall study. J.A.H., H.S., H.VH., C.O., L.O., X.G., A.C., and C.M.R. performed and interpreted computational analyses. X.G. performed *in vitro* experiments. X.G., N.W., A.Q., D.C., A.C., D.F.C., A.J.C., T.C., G.K.F., A.A.R., A.R., J.T., K.H., N.F.K., M.F.K., D.J.E., K.K., A.S., Z.L., C.S.C., P.G.W., R.G., E.M., A.B., B.S.Z., C.L., C.M.H., M.G.P.vdW, G.C.H., T.G., R.B., D.S.L., J.R.G., Y.S., R.P., A.O., A.W., C.J.Y., UCSF COMET Consortium, T.R., J.M.M., F.C., A.T., M.B., L.T., I.O.R., A.R., S.B.K., R.N.B., C.M.R. facilitiated the COMET or COVACTA studies and providing data and/or critical feedback on methods and results. C.M.R., J.A.H, and H.S. wrote the first draft of the manuscript. J.A.H is listed first for defining the ENRAGE signature and responsible for analysis of COMET and public datasets, with H.S. responsible for analysis of the COVACTA study. All authors reviewed and approved the final manuscript.

## Acknowledgements

This study was supported with funding from Roche, Inc. and federal funds from the Department of Health and Human Services; Office of the Assistant Secretary for Preparedness and Response; Biomedical Advanced Research and Development Authority, under OT number: HHSO100201800036C. C.J.Y. is further supported by the NIH grants R01AR071522, R01AI136972, U01HG012192, and the Chan Zuckerberg Initiative, and is an investigator at the Chan Zuckerberg Biohub and is a member of the Parker Institute for Cancer Immunotherapy (PICI). G.C.H. was supported by the National Science Foundation Undergraduate Research Fellowship Program 1650113. C.S.C. is further supported by NIH R35HL140026. C.H. is further supported by a K23 from NHLBI K23 HL133495.

UCSF COMET Consortium co-authors: Yumiko Abe-Jones, Michael Adkisson, K. Mark Ansel, Saurabh Asthana, Alexander Beagle, Sharvari Bhide, Cathy Cai, Saharai Caldera, Maria Calvo, Sidney A. Carrillo, Suzanna Chak, Stephanie Christenson, Zachary Collins, Spyros Darmanis, Angela Detweiler, Catherine DeVoe, Walter Eckalbar, Jeremy Giberson, Ana Gonzalez, Gracie Gordon, Paula Hayakawa Serpa, Alejandra Jauregui, Chayse Jones, Serena Ke, Divya Kushnoor, Tasha Lea, Deanna Lee, Aleksandra Leligdowicz, Yale Liu, Salman Mahboob, Lenka Maliskova, Michael Matthay, Elizabeth McCarthy, Priscila Muñoz-Sandoval, Norma Neff, Viet Nguyen, Nishita Nigam, Randy Parada, Maira Phelps, Logan Pierce, Priya Prasad, Sadeed Rashid, Gabriella Reeder, Nicklaus Rodriguez, Bushra Samad, Andrew Schroeder, Cole Shaw, Alan Shen, Austin Sigman, Pratik Sinha, Matthew Spitzer, Sara Sunshine, Kevin Tang, Luz Torres Altamirano, Alexandra Tsitsiklis, Erden Tumurbaatar, Vaibhav Upadhyay, Alexander Whatley, Andrew Willmore, Michael Wilson, Juliane Winkler, Kristine Wong, Kimberly Yee, Michelle Yu, Mingyue Zhou, Wandi S. Zhu

