## Supplemental Data for "A myeloid program associated with COVID-19 severity is decreased by therapeutic blockade of IL-6 signaling"

**Supplemental Table 1.** EN-RAGE gene set

| ID | Gene symbol | ID | Gene symbol | ID | Gene symbol | ID | Gene symbol |
|---|---|---|---|---|---|---|---|
| ENSG00000276900 | AC023157.3 | ENSG00000163412 | EIF4E3 | ENSG00000248323 | LUCAT1 | ENSG00000101236 | RNF24 |
| ENSG00000268734 | AC245128.3 | ENSG00000198734 | F5 | ENSG00000185022 | MAFF | ENSG00000163221 | S100A12 |
| ENSG00000233461 | AL445524.1 | ENSG00000186431 | FCAR | ENSG00000125505 | MBOAT7 | ENSG00000143546 | S100A8 |
| ENSG00000161944 | ASGR2 | ENSG00000085265 | FCN1 | ENSG00000140563 | MCTP2 | ENSG00000163220 | S100A9 |
| ENSG00000156127 | BATF | ENSG00000125740 | FOSB | ENSG00000257335 | MGAM | ENSG00000213694 | S1PR3 |
| ENSG00000113916 | BCL6 | ENSG00000171051 | FPR1 | ENSG00000172965 | MIR4435-2HG | ENSG00000155307 | SAMSN1 |
| ENSG00000163823 | CCR1 | ENSG00000171049 | FPR2 | ENSG00000008516 | MMP25 | ENSG00000197632 | SERPINB2 |
| ENSG00000125810 | CD93 | ENSG00000123689 | G0S2 | ENSG00000059728 | MXD1 | ENSG00000197208 | SLC22A4 |
| ENSG00000158825 | CDA | ENSG00000151948 | GLT1D1 | ENSG00000105835 | NAMPT | ENSG00000147454 | SLC25A37 |
| ENSG00000131873 | CHSY1 | ENSG00000170837 | GPR27 | ENSG00000165030 | NFIL3 | ENSG00000059804 | SLC2A3 |
| ENSG00000136026 | CKAP4 | ENSG00000139572 | GPR84 | ENSG00000100906 | NFKBIA | ENSG00000082014 | SMARCD3 |
| ENSG00000166527 | CLEC4D | ENSG00000136630 | HLX | ENSG00000087157 | PGS1 | ENSG00000122862 | SRGN |
| ENSG00000166523 | CLEC4E | ENSG00000173083 | HPSE | ENSG00000105520 | PLPPR2 | ENSG00000180953 | ST20 |
| ENSG00000120885 | CLU | ENSG00000160888 | IER2 | ENSG00000163421 | PROK2 | ENSG00000010327 | STAB1 |
| ENSG00000146592 | CREB5 | ENSG00000137331 | IER3 | ENSG00000140368 | PSTPIP1 | ENSG00000127954 | STEAP4 |
| ENSG00000103196 | CRISPLD2 | ENSG00000115594 | IL1R1 | ENSG00000125384 | PTGER2 | ENSG00000166900 | STX3 |
| ENSG00000163739 | CXCL1 | ENSG00000136689 | IL1RN | ENSG00000073756 | PTGS2 | ENSG00000137462 | TLR2 |
| ENSG00000169429 | CXCL8 | ENSG00000157551 | KCNJ15 | ENSG00000155093 | PTPRN2 | ENSG00000173334 | TRIB1 |
| ENSG00000138061 | CYP1B1 | ENSG00000239998 | LILRA2 | ENSG00000089159 | PXN | ENSG00000127824 | TUBA4A |
| ENSG00000139318 | DUSP6 | ENSG00000187116 | LILRA5 | ENSG00000105514 | RAB3D | ENSG00000038427 | VCAN |
| ENSG00000135636 | DYSF | ENSG00000182541 | LIMK2 | ENSG00000169385 | RNASE2 | ENSG00000229124 | VIM-AS1 |

**Supplementary Table 2.** Reproducibility of EN-RAGE+ myeloid cell gene expression across cohorts and sample types. Spearman correlation coefficients are shown for pseudo-bulk EN-RAGE signature score and genes encoding myeloid functions within specific myeloid populations across scRNA-seq datasets. Positive correlations are shaded red and negative correlations shaded blue, with increasing darkness of shading indicating two tailed p values of $p<0.05$, $p<0.01$, and $p<0.001$. TA=tracheal aspirate, BAL=bronchoalveolar lavage. Blanks indicate transcript in below detection. Datasets: Liao et al.[7], Grant et al.[6], Delorey et al.[5], Schulte-Schrepping et al.[11], Silvin et al.[12], COMET PBMC[2], whole blood[37], and ETA[36].

| | Spearman rho | | HLA.DRA | HLA.DRB1 | CD14 | STAT3 | S100A12 | S100A9 | IL10 | PDL1 | PTGER2 | TGFB1 | IL1B | IDO1 | CCR5 | CYBB | ARG1 | CCR2 |
|---|---|---|---|---|---|---|---|---|---|---|---|---|---|---|---|---|---|---|
| Airway samples | **Monocyte** | n | | | | | | | | | | | | | | | | |
| | TA MonoMac COMET | 41 | 0.11 | -0.23 | 0.10 | 0.75 | 0.70 | 0.76 | 0.74 | 0.59 | 0.72 | 0.13 | 0.86 | 0.33 | 0.36 | 0.14 | 0.31 | 0.26 |
| | BAL monocyte Liao | 12 | -0.90 | -0.97 | 0.73 | 0.71 | 0.92 | 0.38 | 0.82 | 0.58 | 0.89 | 0.89 | 0.74 | 0.69 | 0.80 | 0.07 | | 0.73 |
| | BAL myeloid Grant | 19 | 0.52 | 0.43 | 0.41 | 0.48 | 0.53 | 0.16 | 0.36 | 0.47 | 0.66 | 0.29 | 0.47 | 0.34 | 0.37 | 0.06 | | -0.05 |
| | BAL macrophage Grant | 19 | -0.26 | -0.33 | 0.32 | 0.83 | 0.93 | 0.89 | 0.65 | 0.76 | 0.81 | 0.38 | 0.55 | 0.61 | 0.46 | -0.19 | 0.30 | 0.15 |
| | Lung myeloid Delorey | 24 | -0.01 | 0.00 | -0.02 | 0.65 | 0.76 | 0.32 | 0.77 | 0.85 | 0.74 | 0.29 | 0.87 | 0.65 | 0.43 | -0.15 | 0.53 | 0.28 |
| | **Neutrophil** | | | | | | | | | | | | | | | | | |
| | TA neutrophil COMET | 40 | 0.05 | -0.16 | 0.58 | 0.63 | 0.74 | 0.85 | 0.53 | 0.61 | 0.35 | 0.54 | 0.78 | 0.30 | -0.02 | 0.21 | 0.59 | -0.09 |
| | BAL neutrophil Liao | 11 | 0.34 | 0.35 | 0.82 | 0.84 | 0.89 | 0.96 | | 0.99 | 0.87 | 0.96 | 0.83 | 0.93 | 0.80 | 0.85 | | 0.15 |

| | Spearman rho | | HLA.DRA | HLA.DRB1 | CD14 | STAT3 | S100A12 | S100A9 | IL10 | PDL1 | PTGER2 | TGFB1 | IL1B | IDO1 | CCR5 | CYBB | ARG1 | CCR2 |
|---|---|---|---|---|---|---|---|---|---|---|---|---|---|---|---|---|---|---|
| Blood samples | **Monocyte** | n | | | | | | | | | | | | | | | | |
| | Blood monocyte COMET | 18 | -0.01 | 0.19 | 0.62 | 0.79 | 0.67 | 0.74 | 0.57 | 0.31 | 0.52 | 0.37 | 0.34 | 0.37 | 0.07 | 0.60 | 0.43 | 0.53 |
| | Blood monocyte Schulte | 33 | -0.54 | -0.41 | 0.81 | 0.75 | 0.67 | 0.67 | 0.05 | 0.27 | 0.51 | 0.36 | -0.16 | 0.09 | 0.05 | 0.56 | -0.01 | 0.23 |
| | Blood monocyte Silvin | 6 | -0.77 | -0.89 | 0.94 | 0.54 | 1.00 | 0.77 | | 0.68 | -0.03 | 0.31 | -0.43 | | 0.43 | 0.94 | | 0.60 |
| | PBMC monocyte COMET | 44 | -0.45 | -0.48 | 0.65 | 0.15 | 0.73 | 0.79 | 0.15 | 0.02 | 0.38 | 0.18 | 0.34 | -0.08 | -0.44 | 0.52 | -0.09 | 0.25 |
| | PBMC monocyte Schulte | 16 | -0.73 | -0.54 | 0.75 | 0.86 | 0.57 | 0.71 | 0.66 | 0.43 | 0.77 | -0.20 | 0.64 | 0.02 | -0.40 | 0.40 | 0.65 | -0.56 |
| | **Neutrophil** | | | | | | | | | | | | | | | | | |
| | Blood neutrophil COMET | 18 | -0.53 | -0.52 | 0.78 | 0.94 | 0.72 | 0.87 | 0.30 | 0.78 | -0.48 | 0.91 | 0.70 | 0.37 | 0.06 | 0.28 | 0.77 | -0.21 |
| | Blood neutrophil Schulte | 33 | -0.23 | -0.05 | 0.71 | 0.79 | 0.57 | 0.62 | | 0.50 | 0.22 | 0.80 | -0.08 | | 0.33 | 0.19 | 0.19 | -0.01 |
| | Blood neutrophil Silvin | 6 | -0.37 | -0.77 | 1.00 | 0.20 | 1.00 | 0.94 | | 0.60 | 0.82 | 0.60 | -0.03 | | | -0.09 | 0.54 | -0.85 |
| | PBMC neutrophil Schulte | 16 | 0.21 | 0.46 | 0.54 | 0.74 | 0.63 | 0.68 | | 0.62 | -0.11 | 0.91 | 0.80 | 0.55 | | 0.23 | 0.47 | |

**Supplementary Figure 1.** Further characterization of the signature within PBMC and BAL. UMAP plots from PBMC and BAL.
A. UMAP projection of cells colored by cell types. Each point represents a single cell, and points are colored by annotated cell types. PBMC[11] or BAL[7].
B. UMAP projection of cells colored by EN-RAGE signature score. Each point represents a single cell, colored by the signature score value.
C. Extended pseudo-bulk expression profiles including non-myeloid cell types. Each point represents the pseudo-bulk signature score for a cell type in a patient sample. Blue=healthy (BAL n = 3; PBMC, n = 3), orange=moderate (BAL, n = 3; PBMC, n = 8; hospitalized +/- supplemental $O_2$), red=critical (BAL, n = 6; PBMC, n = 10; requiring mechanical ventilation), with severity defined within each published dataset by the original authors.
D-G. Heatmaps of ENRAGE (D-E) and MS1 (F-G) signature genes in pseudo-bulk expression profiles in BAL and PBMC. Each row is a gene, each column is a cell cluster. Each row is z-score normalized within each dataset. LDG=low density granulocytes.

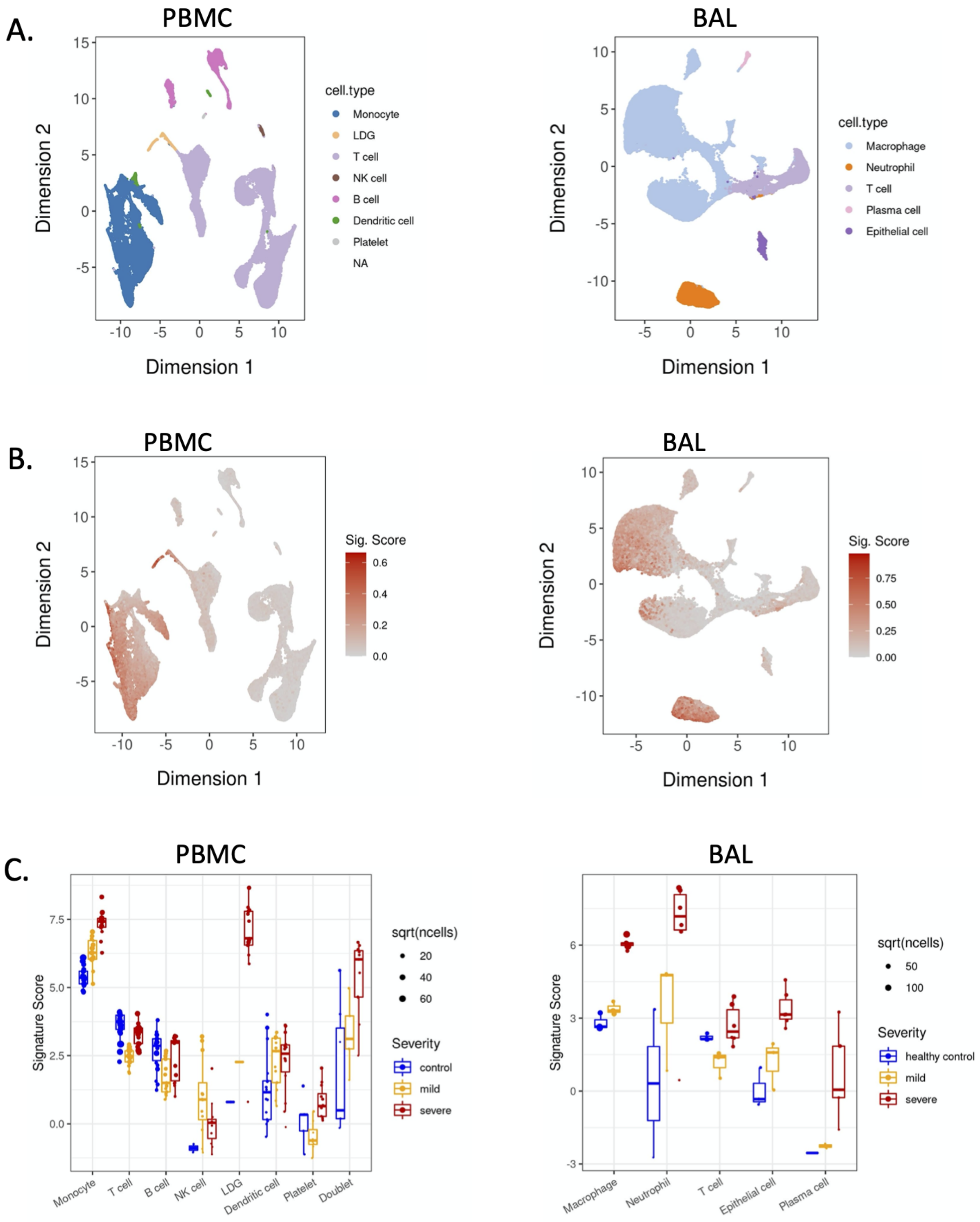
A.
PBMC
BAL
cell.type
Monocyte
LDG
T cell
NK cell
B cell
Dendritic cell
Platelet
NA
Macrophage
Neutrophil
Plasma cell
Epithelial cell
Dimension 1
Dimension 2
B.
PBMC
BAL
Sig. Score
C.
PBMC
BAL
Signature Score
sqrt(ncells)
Severity
control
mild
severe
healthy control
Doublet

**Supplementary Figure 2.** Performance of ENRAGE signature across sample types (COMET cohort). **A.** Pairwise Spearman correlation coefficients between genes within the ENRAGE and MS1 gene signatures across three different compartments: ETA (n = 55), PBMC (n = 279), and whole blood (n = 245). **B-E.** Heatmaps of gene expression (B-C) and pairwise gene correlations (D-E) of ENRAGE and MS1 within the PBMC compartment. (11). **F-I.** Heatmaps of gene expression (F-G) and pairwise gene correlations (H-I) of the whole blood compartment (11). **J-M.** Heatmaps of gene expression (J-K) and pairwise gene correlations (L-M) of ENRAGE and MS1 within the ETA compartment (7). Gene expression values were pseudobulked for each cell population (sample + cell type) and the pairwise Spearman correlation coefficients were calculated using the pseudobulked gene expression values. LDG=low-density granulocytes.

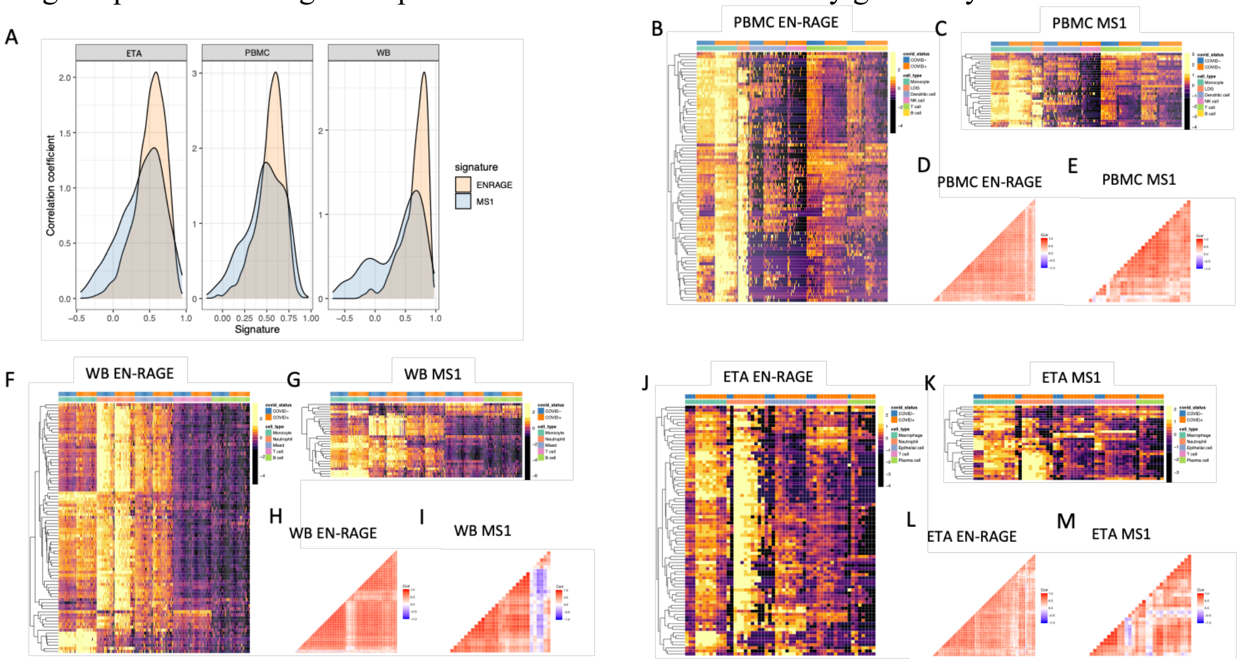

**Supplementary Figure 3.** EN-RAGE gene set expression in PBMCs is associated with increased clinical severity and poor outcomes in the COMET cohort. **A.** Comparison of EN-RAGE signature in monocytes from patients admitted to ICU compared to those not. **B-C.** Correlation between baseline pseudobulk EN-RAGE gene set expression in myeloid cells and B. baseline NIH scale, and C. maximal NIH scale (worst score recorded during hospitalization, excluding healthy controls). Each dot represents one patient, n=49. D-F. Myeloid EN-RAGE gene set expression by ARDS diagnosis. **D.** ARDS diagnosed using AECC criteria. **E.** ARDS diagnosed using Berlin criteria. **F.** Berlin ARDS subset by COVID-19 status. Boxes represent first and third quartiles, the centerline shows the median. Two-tailed t-test p-values are shown: n.s.= not significant, *p<0.05, **p<0.01.

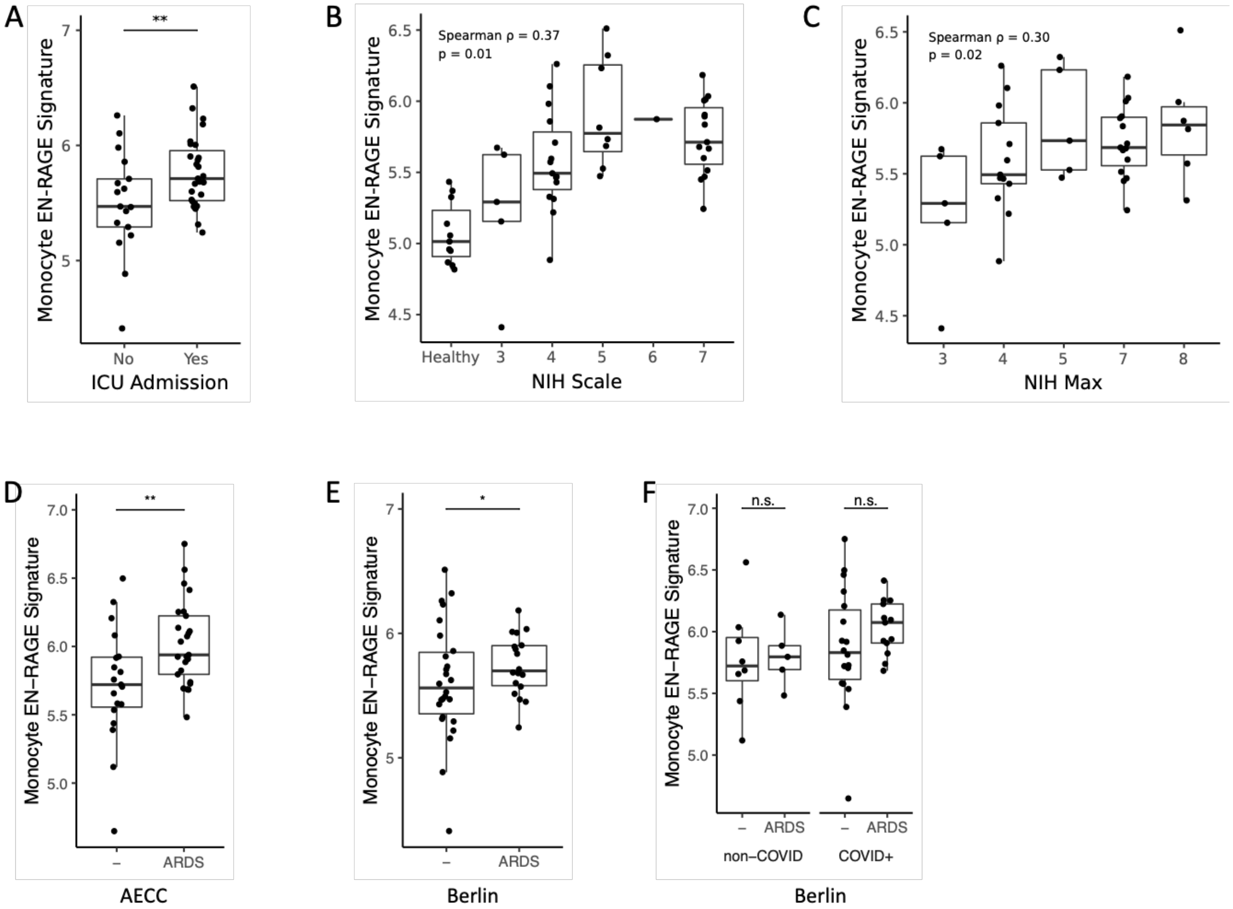

**Supplemental Figure 4.** Characterization of T cell immunosuppression phenotypes (COMET PBMC cohort; 128 samples from 60 patients [429, 505 cells]).
Pseudobulked surface protein expression in 128 PBMC samples from 60 patients over 14 days in patients grouped by clinical outcomes. A-D. $CD8^+$ T cells. E-H. $CD4^+$ T cells. Blue line denotes the linear regression trend for gene expression over time. Red line denotes the average expression level in 11 healthy controls. Vent. duration = days of mechanical ventilation in survivors. Pearson correlation coefficients (r) and p values are indicated.

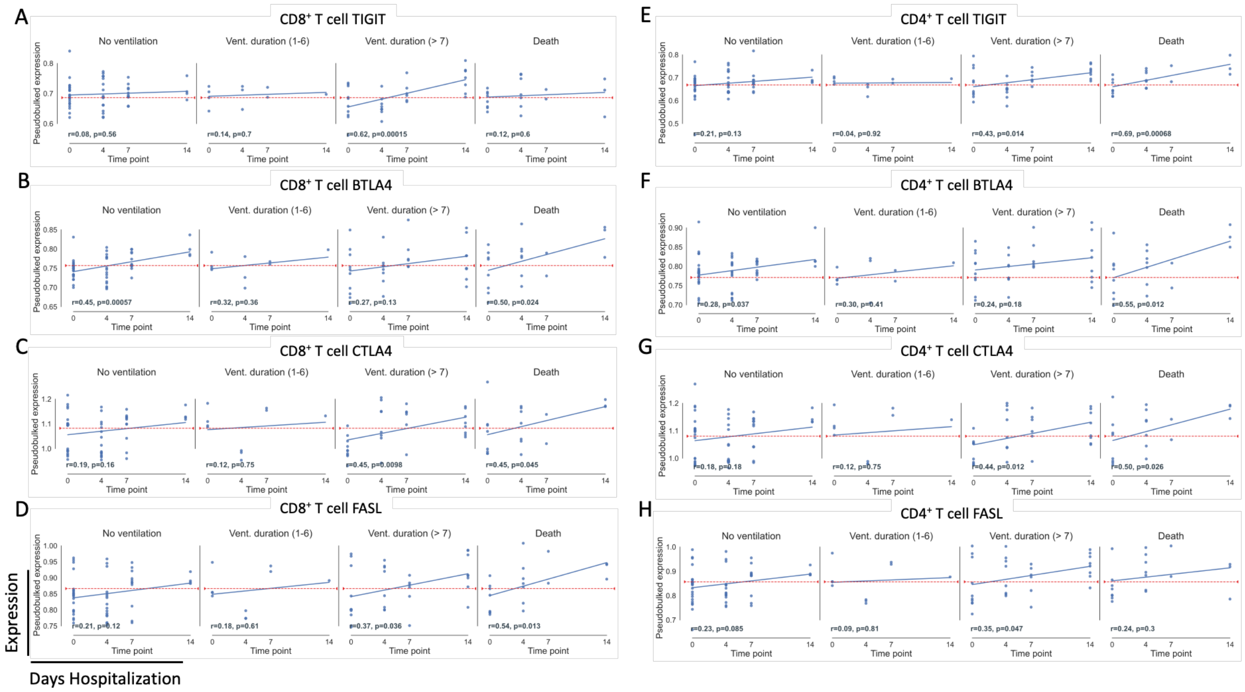

**Supplemental Figure 5.** Myeloid and T cell gene sets are associated with poor outcome in COVID-19 patients and decreased by tocilizumab treatment.
GSEA for EN-RAGE and MS1 gene sets are similar. GSEA enrichments are consistent with those in Figure 6B-E following adjustment for A. baseline blood % monocytes, B. baseline % neutrophils, or C. baseline % lymphocytes. D-E. The effect of tocilizumab treatment is not affected by baseline tertile of serum IL-6 protein levels, with 1= lowest tertile and 3=highest tertile. GSEA are shown when FDR<0.05 (red) and shaded grey when p<0.01 but FDR>0.05. NES=normalized enrichment score.

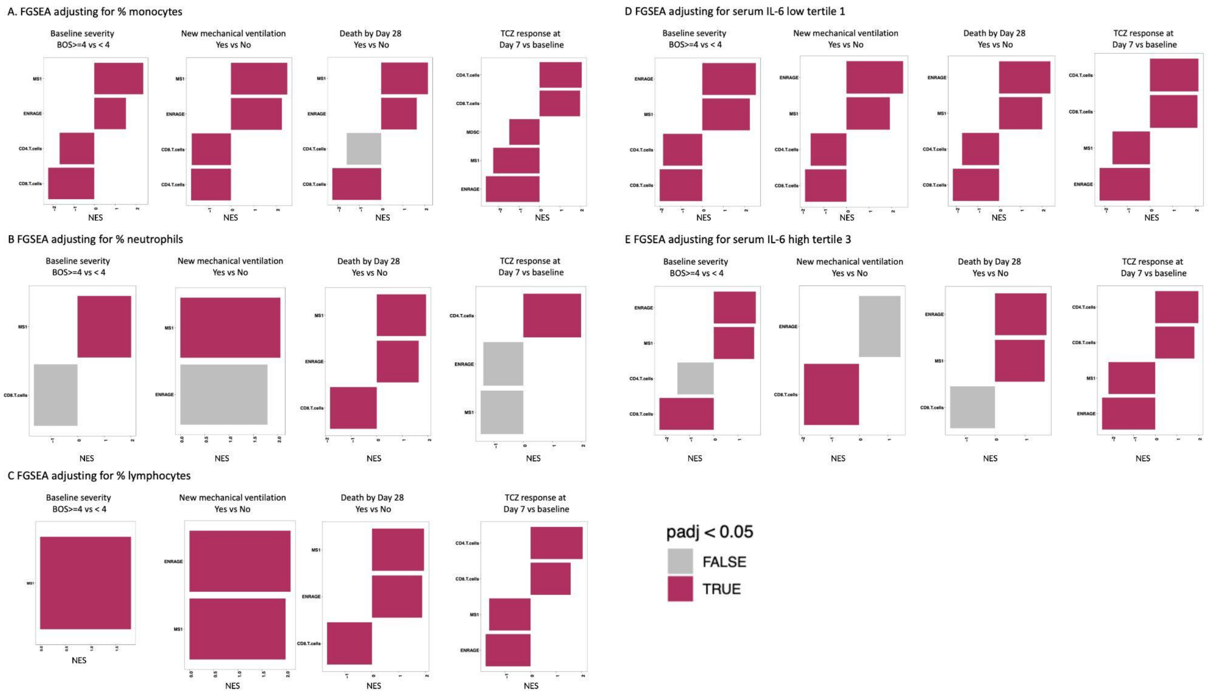

**Supplemental Figure 6. A-D.** Tocilizumab increases expression of blood myeloid and T cell signature genes and cell composition more than placebo over 7 days of treatment. Signature scores are shown within each day, split by treatment arm and disease course and compared with healthy controls (CTRL). Patients are split into those that were discharged before D28, those who remained hospitalized, or subjects who died by day 28. Average signature scores are shown across the first 7 days of treatment for patients with measurements at all 3 time points. **A-B**. Tocilizumab treatment decreases expression of **(A)** ENRAGE and **(B)** MS1 myeloid signatures in survivors. **C-D.** Tocilizumab treatment increased expression of **(C)** CD8$^{+}$ and **(D)** CD4$^{+}$ T cell signatures in survivors. **E-J**. Tocilizumab normalizes blood cell composition more rapidly than placebo. Blood cell percentage (**E-G**) and absolute cell counts (**H-J**) are shown at D1 (pre-treatment) and D3 and D4-D10 (post-treatment); FDR calculated relative to D1 * = $p < 0.05$, ** = $p < 0.01$, *** = $p < 0.001$, **** = $p < 0.0001$, ns = non-significant. Significance testing was performed using either t-test (A-D) or Wilcoxon rank-sum test (E-J). SOC = standard of care drug therapy. For box plots, each point represents a patient sample, center lines depict the median value, the bottom and top of the boxes show the first and third quartile, and whiskers show the most extreme point <1.5x the interquartile range (IQR) from the bottom or top of the box.

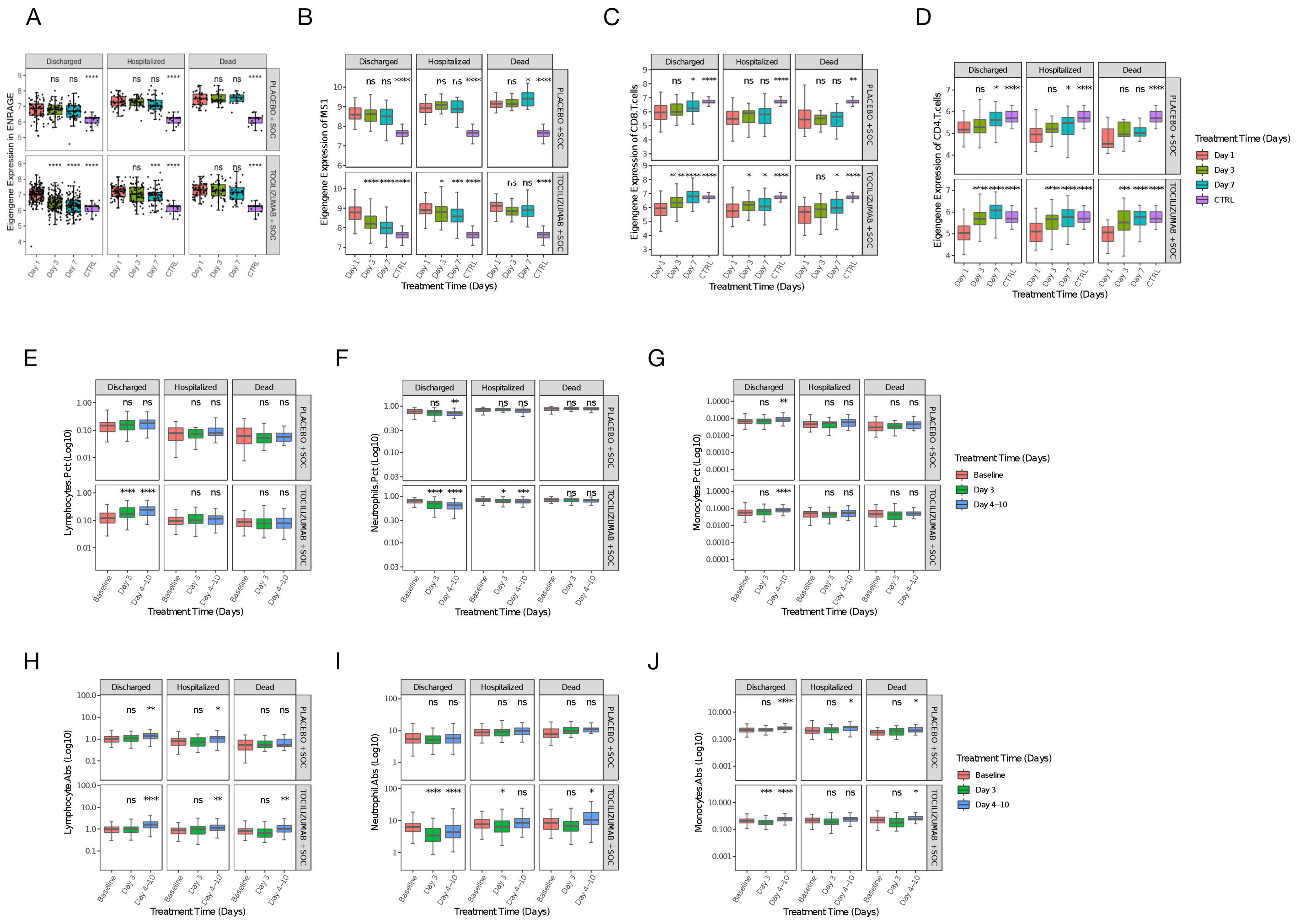

**Supplemental Figure 7.** Effect of tociluzimab on blood cell counts and serum protein levels relative to ENRAGE gene set expression Day 7 vs Day 1. A. Correlation of blood ENRAGE gene set expression with serum proteins and blood cell counts on D1 prior to treatment. IL-10 was measured by Protein Simple and IL-1β, ENRAGE, and Arg1 were measured by Olink. D-H. Spearman correlations between change in blood ENRAGE gene set expression and change in blood cell counts (B-D) and serum proteins (E-H) between D1 and D7. Each dot represents a patient with available samples at D1 and D7, SOC = standard of care drug therapy.

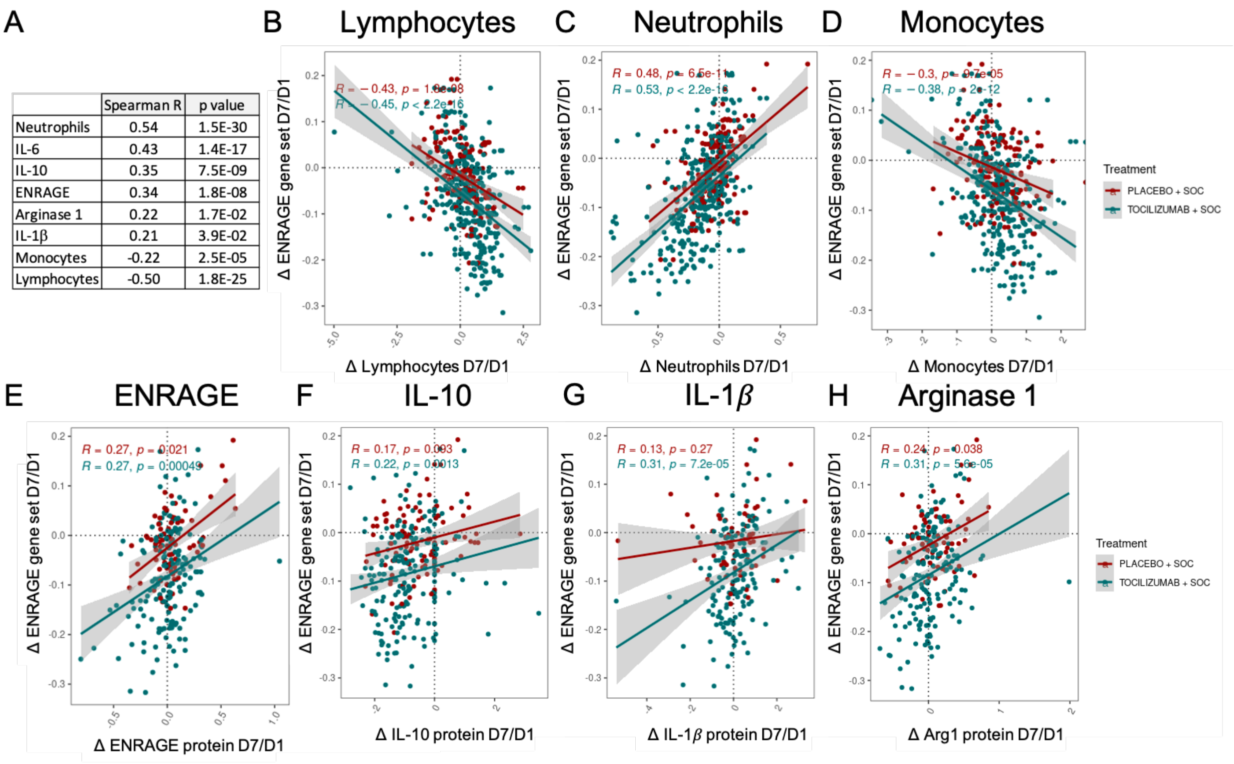


| | Spearman R | p value |
|---|---|---|
| Neutrophils | 0.54 | 1.5E-30 |
| IL-6 | 0.43 | 1.4E-17 |
| IL-10 | 0.35 | 7.5E-09 |
| ENRAGE | 0.34 | 1.8E-08 |
| Arginase 1 | 0.22 | 1.7E-02 |
| IL-1β | 0.21 | 3.9E-02 |
| Monocytes | -0.22 | 2.5E-05 |
| Lymphocytes | -0.50 | 1.8E-25 |
